# Alzheimer’s Disease variant portal (ADVP): a catalog of genetic findings for Alzheimer’s Disease

**DOI:** 10.1101/2020.09.29.20203950

**Authors:** Pavel P. Kuksa, Chia-Lun Liu, Wei Fu, Liming Qu, Yi Zhao, Zivadin Katanic, Amanda B Kuzma, Pei-Chuan Ho, Kai-Teh Tzeng, Otto Valladares, Shin-Yi Chou, Adam C Naj, Gerard D Schellenberg, Li-San Wang, Yuk Yee Leung

## Abstract

Alzheimer’s Disease (AD) genetics has made substantial progress through genome-wide association studies (GWASs). An up-to-date resource providing harmonized, searchable information on AD genetic variants with linking to genes and supporting functional evidence is needed.

We developed the Alzheimer’s Disease Variant Portal (ADVP), an extensive collection of associations curated from >200 GWAS publications from Alzheimer’s Disease Genetics Consortium (ADGC) and other researchers. Publications are reviewed systematically to extract top associations for harmonization and genomic annotation.

ADVP V1.0 catalogs 6,990 associations with disease-risk, expression quantitative traits, endophenotypes and neuropathology across >900 loci, >1,800 variants, >80 cohorts, and 8 populations. ADVP integrates with NIAGADS Alzheimer’s GenomicsDB where investigators can cross-reference other functional evidence.

ADVP is a valuable resource for investigators to quickly and systematically explore high-confidence AD genetic findings and provides insights into population- and tissue-specific AD genetic architecture. ADVP is continually maintained and enhanced by NIAGADS and is freely accessible (https://advp.niagads.org).

## 1. Introduction

Alzheimer’s Disease (AD) is a devastating neurological disorder affecting millions of people worldwide and is the most common cause of dementia [1]. There are no approved drugs that can slow or treat the disease. The disease is complex and highly heritable [2]. The strongest known genetic risk factor for AD is the ε4 allele of the Apolipoprotein E gene (*APOE* ε4) [3, 4], but more than one-third of AD cases do not carry any *APOE* ε4 alleles. Large-scale genome-wide association studies (GWASs) have led to the discovery of additional common genetic loci associated with the late-onset AD (LOAD) [5–9]. Yet, the identification of genetic contributors to LOAD remains a challenge as LOAD is likely caused by multiple low penetrance genetic variants [10], with the small sample sizes further complicating the identification of these causal variants.

The Alzheimer’s Disease Genetics Consortium (ADGC) was founded in 2009 and funded by National Institute on Aging (NIA), to conduct large sample GWAS to identify genes associated with an increased risk of developing LOAD. ADGC co-founded IGAP (International Genomics of Alzheimer’s Project) with three other AD genetics consortia: Cohorts for Heart and Aging Research in Genomic Epidemiology (CHARGE) Consortium, the European Alzheimer’s Disease Initiative (EADI), and the Genetic and Environmental Risk in Alzheimer’s Disease (GERAD) Consortium. IGAP assembled large Caucasian samples for better statistical power and was able to identify 19 genome-wide significant loci in 2013 [11], and five more loci using more than 30,000 samples in 2019 [12].

In addition to GWA studies focused on association with disease risk, recently many genetics studies have focused on related phenotypes including neuroimaging biomarkers [13], circulating biomarkers in [14, 15], cognitive decline [16, 17] neuropathology [18], family history [19]. GWAS on Hispanic, African-American, Asian, and other minority populations also led to new variants not observed in Caucasians [20–22] In order to help investigators better explore the rich and diverse literature of genetic findings, it is important to have a single resource with harmonized, unified, searchable information on identified genetic variants and genes across a variety of AD studies and populations, along with supporting functional genomic evidence.

To meet this need, we have cataloged genetic association results (both genome-wide significant associations [*p*≤5×10^-8^ by convention] and all other associations highlighted in the main text from all major GWA studies published by ADGC from 2009 to 2019 and other AD GWAS articles identified from the NHGRI/EBI GWAS Catalog (Buniello et al., 2019). Summaries extracted from each of the articles are made publicly available on a continuously updated and freely accessible Alzheimer’s Disease Variant Portal (ADVP) (https://advp.niagads.org). To date, ADVP provides the largest, most updated, and comprehensive collection of systematically curated, harmonized, and annotated AD-specific genetic associations. This first release contains information on 6,990 genetic associations, >900 genomic loci curated from >125 AD publications categorized into nine harmonized phenotype categories. All AD associations in ADVP are annotated with genomic and functional information. Comprehensive biological annotations are available via integration with the NIAGADS Alzheimer’s Disease Genomics database [24]. ADVP will serve as an invaluable resource for the research community to explore and decipher the genetic architecture of AD and other neurodegenerative diseases.

## 2. Methods

An overview of the ADVP study design is shown in Figure 1.

**Figure 1.**
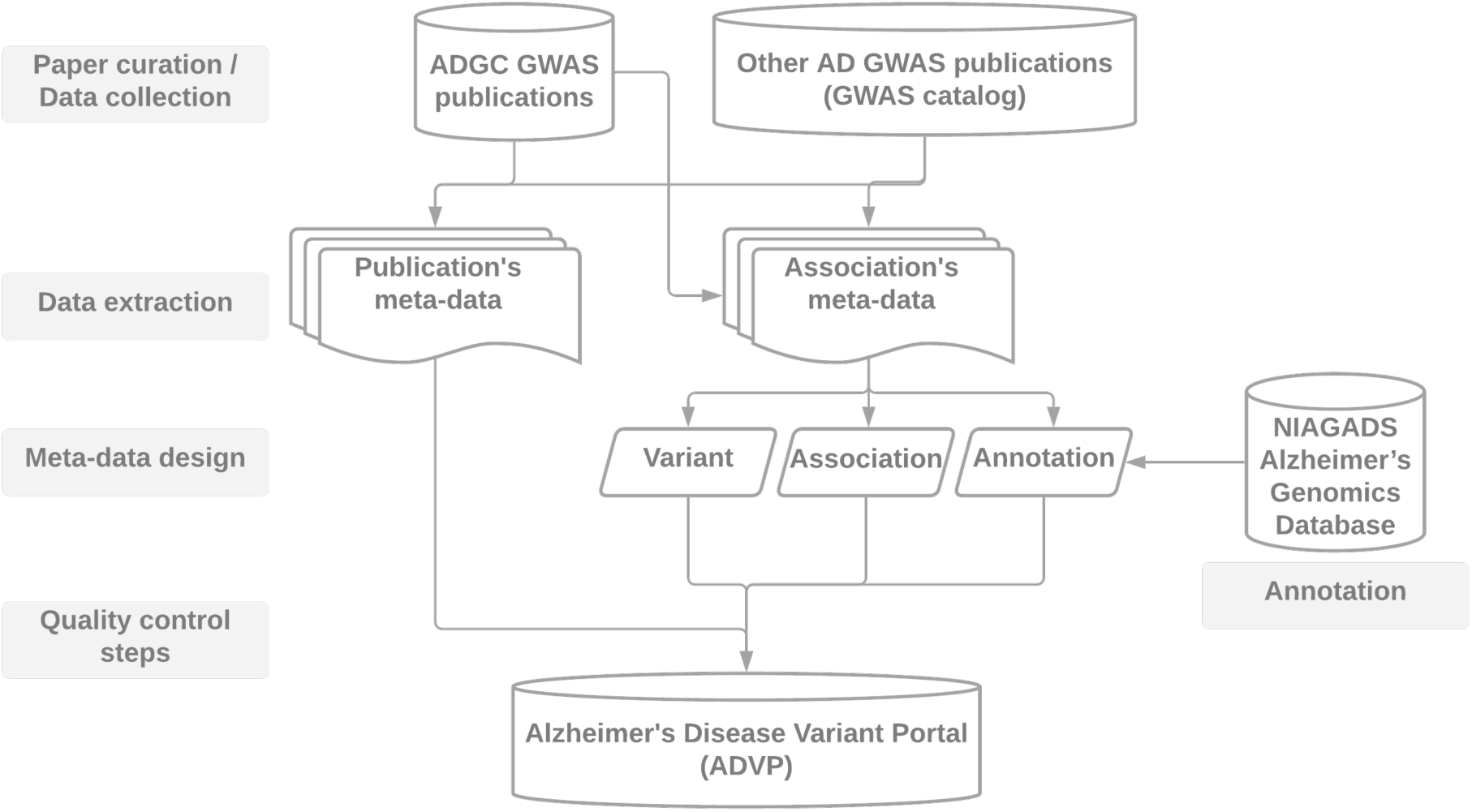
ADVP study design. AD GWAS publications are first collected (Section **“2.1 Data collection”**), genetic variant and association data are then systematically extracted (Section “**2.2 Data extraction**”), harmonized (Section “**2.3 Meta-data design**”), annotated (Section “**2.4 Annotation**”), subjected to quality control steps (Section “**2.5 Quality control steps**”) and stored into ADVP.

### 2.1 Collection and curation of AD-related GWAS publications (Data collection)

This ADVP V1.0 release consists of curated and harmonized genetic associations from the genome-wide significant and suggestive loci collected from AD genetic studies conducted primarily by the ADGC. All AD GWAS publications by ADGC (2009-2019, http://www.adgenetics.org) and all other AD GWA studies in GWAS catalog [23] (MeSH D000544, curation date: Dec 2019) were included. All the publications (total N=205; ADGC: N=134; Citations from ADGC: N=20; GWAS catalog: N=51) were first screened to identify publications reporting GWAS findings. For each publication, all main genetic associations reported in the main text (table format) were systematically extracted. As the current ADVP release focuses on the major/main findings in each publication/GWA study, we did not extract associations reported in the supplementary tables as they often represent supplemental findings not reaching the same kind of statistical significance as the main associations. In total, we curated the 125 publications which contain GWAS findings in the main text (https://advp.niagads.org/publications). **Supplementary Table S1** provides details on all curated AD publications in ADVP V1.0. Note the ADGC family-based analyses results will be included in the next release.

### 2.2 Extraction of genetic variants and associations from publications (Data extraction)

We applied the following systematic data extraction and curation procedure for each publication to organize all the extracted variant and association information into a structured tabular format according to the ADVP data schema (see Section “**2.3 Meta-data design**” for details about the columns). In each publication, we identified all the tables in the main text with reported association *p*-values. All the information for these associations was then saved into a standardized template document using the corresponding meta-data schema and the two predefined worksheets: publication meta-data and association meta-data. Lastly, the document is parsed by customized scripts to normalize, validate, annotate, and store the publication, variant, and association data in the relational database [33]. Collected AD variants and association records are further linked (using positional and variant and gene identifier information) with the NIAGADS Alzheimer’s Genomics Database [24] which provides the comprehensive annotation and functional genomic information. **Supplementary Note 2** provides details on ADVP architecture and implementation.

### 2.3 Meta-data schema for systematic curation and harmonization of genetic associations (Meta-data design)

#### 2.3.1 Publication meta-data

Meta-data for all curated publications in ADVP was extracted from PubMed (https://pubmed.ncbi.nlm.nih.gov) using the NCBI EDirect service (https://www.ncbi.nlm.nih.gov/books/NBK179288) with publication PubMed identifiers (PMID) as query keys. For each publication, we record its PMID, PubMed Central identifier (PMCID), first and last authors, journal, and year of publications. We also store the abstract, article URLs, and information on curated/source tables in the Publication meta-data (Figure 1).

#### 2.3.2 Association meta-data

ADVP association meta-data consists of 28 data fields, of which 19 are extracted directly from the paper contents. The rest are harmonized (based on extracted original information) and programmatically generated fields. Altogether, association meta-data provide 1) variant information (Section “**2.3.2.1 Description of Variants”**); 2) association information (Section “**2.3.2.2 Description of association records”**); and 3) annotation information (Section “**2.4 Annotation of genetic variants and associations”**). For a detailed explanation of these curated and harmonized/derived data fields, see **Supplementary Table S2**.

#### 2.3.2.1 Description of Variants

Each genetic variant in ADVP is described using dbSNP rsID, genomic coordinates (chromosome:basepair), reference and alternative alleles. Both the information reported in the publication (if available) and those derived from the reference databases such as dbSNP [25] and NIAGADS Alzheimer’s Genomics database [24], are included in the variant description. Genomic location in ADVP is currently stored using GRCh37/hg19 reference genome build as the majority of GWAS publications conducted analyses using GRCh37/hg19.

#### 2.3.2.2 Description of association records

The primary association information was systematically extracted from each source table and recorded as part of the ADVP association record. The extracted information was further recoded and categorized, so that association records are described consistently across publications. For each reported association we first collected a pre-defined set of data attributes commonly reported by genetic association studies (See “Extracted” columns under **Supplementary Table S2**). These include *p*-value and statistics related to the effect size (regression beta coefficients and variance, odds ratios, confidence intervals), reported effect allele and its frequency in the studied population.

In addition to the information directly extracted from publications, each association in ADVP is described with the nine harmonized meta-information data fields (see **Supplementary Note 1** for more details on each of these fields; field names are denoted with the double quotes below):

1. “Record type”: association record type.
2. “Population”: normalized study population information.
3. “Cohort”: normalized cohort names.
4. “Sample size”: original sample size.
5. “Subset analyzed”: description of the subset of samples used to perform the association analysis.
6. “Phenotype”: the outcome variable (phenotype/trait) of the association analysis.
7. “Association Type”: standardized type of association (disease-risk, eQTL, endophenotype, and others).
8. “Stage”: analysis stage (discovery, validation, replication, or meta- or joint-analysis). Please see (**Supplementary Figure 1, Supplementary Note 1**) for details on how this information was derived.
9. “Imputation”: imputation panel information.

Note, “Population”, “Cohort”, and “Phenotype” information are displayed in ADVP using both the original (reported) and the derived, harmonized data fields.

### 2.4 Functional genomics evidence for genetic variants and associations (Annotation)

All variants and association in ADVP were systematically annotated with genomic context (closest upstream/downstream genes), genomic element (promoter, UTR, intron, exon, intergenic, repeat), functional impact (variant most severe consequence), and cross-referenced to NIAGADS Alzheimer’s Genomics database [24].

ADVP reports the genomic context of each genetic variant via multiple data fields: 1) “Locus” – records the gene name as reported in the publication; 2) “Nearest gene” – contains the name of the gene closest to the variant and the distance to the gene (in base pairs (bps)) in upstream (+) or downstream (-) orientation. The nearest genes are identified using GENCODE v34 [26] protein-coding gene annotations. For each genetic variant co-localized with one or more genes, both EnsemblID [27] and HGNC [28] symbols for the gene(s) are reported. For each ADVP variant, the co-localized genomic element is reported based on the genomic partition information [29, 30].

### 2.5 Variant and association data verification (Quality control steps)

Quality control for the variant and association information in ADVP is carried out at multiple levels:

1. We ensured records are not double-counted/re-reported across studies. Each association record in ADVP is uniquely identified by a combination of reported gene/SNP/interaction name, cohort/analyzed subset, association model used, phenotype, and association *p*-value and effect size.
2. We cross-checked recorded positional information (chromosome:basepair), rsID, and allele information against reference databases including, dbSNP [25], NIAGADS Alzheimer’s Genomics database [24], 1000 Genome data [31] to ensure variant information is correct.
3. We identified and removed records solely representing associations annotated by publicly available resources such as GTEx [32].

### 2.6 Population-based analysis of AD associations

To understand the genetic architecture of AD associated loci across populations, we compared their reported effect sizes and frequencies for each locus. In this analysis, we used all association records (from case-control GWAS studies) with complete information on reported allele, effect size (odds ratio), and allele frequency. We then analyzed the four major populations with the most association records (African American, Asian, Caribbean Hispanic, and Caucasian/Non-Hispanic White). Any association records with *p*>0.01 were excluded from analysis. Then for each AD-associated locus (identified using the nearest gene), we selected the association record with the most significant (smallest) *p*-value as a representative record for the locus.

To perform comparison, we normalized the original (reported) odds ratios to reflect the effect of the minor allele. We further categorized the AD-associated loci as population-specific or shared if they were associated with AD in more than one population under study. We then compared AD-associated loci (both shared and population-specific) across populations based on their normalized effect sizes (odds ratios for minor alleles) and the minor allele frequency.

### 2.7 Functional analysis of AD associations

To perform functional analysis of the variants in ADVP, we evaluated the significance of overlaps between ADVP variants and active enhancer elements across tissues and cell types. We used Roadmap Epigenomics [33] (ChromHMM-determined [34]) and FANTOM5 [35] enhancer sets across 35 tissue/cell type categories. We then ranked individual tissue/cell types based on the degree and significance (odds ratio, Fisher’s exact test) of enhancer overlaps in each of the analyzed tissues/cell types.

## 3. Results

In terms of the number of curated AD-related associations and publications, ADVP is more comprehensive than the NHGRI-EBI GWAS catalog [23] (Table 1), the premier general catalog of published GWAS results, and more recent than AlzGene [36], another major database of published AD genetic association studies. In order to focus on association findings with the highest confidence, we decided to concentrate on large-scale association studies at the genomic level, with the majority of studies included in ADVP (65%) reporting associations reaching genome-wide significance, a gold standard for human genetic discoveries. Furthermore, ADVP collected extensive meta-data, including consortiums and cohorts, which were not available in the other two databases and are important for relating the results reported across publications. Finally, ADVP provides convenient links for investigators to explore biological significance of the reported variants (e.g., their genomic context, available functional genomic data or other known associations, if any) via an annotation in NIAGADS Alzheimer’s Genomics database [24] (Figure 1). Following the ADVP curation criteria (see “**Data collection**”, Figure 1), we first identified and screened 205 AD-related publications from 2009-2019. Out of these, we identified 125 publications with genetic associations reported in the main text tables (N=225 tables). Genetic variant and association data were then systematically extracted (Section “**2.2 Data extraction**”), harmonized (“**2.3 Meta-data design**”) (converted into standard variant/association descriptors), annotated (“**2.4 Annotation**”), subjected to quality control steps (“**2.5 Quality control steps**”) and stored in ADVP (Figure 1).

**Table 1:** Comparison between ADVP and existing AD genetics database (AlzGene and GWAS Catalog).

| Features | AlzGene [36] | GWAS Catalog [23] | ADVP |
| --- | --- | --- | --- |
| Focus on AD | Yes | No | Yes |
| Types of genetic associations recorded / included | Variant only | Variant only | Variant, Gene, SNP-SNP and gene-gene interactions |
| Included associations ($p$-value cutoff) | Genome-wide significant only ($<5 \times 10^{-8}$) | Genome-wide significant only ($<5 \times 10^{-8}$) | Both genome-wide significant and suggestive associations |
| Number of curated AD GWAS publications (publication years) | 41 (1998-2011) | 69 (2007-current) | Initial set: 205; Curated set: 125 (2009-current) |
| Number of curated records/associations | 264 genes | 1,532 records (1,155 variants) | 6,605 AD, 6,990 total association records (1,825 variants) |
| Genomic and functional genomic annotations | No | No | NIAGADS Alzheimer's Genomics database [24] annotations |
| Detailed / extensive meta-data | Population, cohort, sample size, phenotype | Population, sample size, stage, phenotype | Record type (SNP/gene-based), population, cohort, sample size, subset analyzed, phenotype, association type, stage, imputation |
| Last update date | 2011 | 2021 | 2021 |

### 3.1 ADVP data summary

The ADVP V1.0 release contains high-quality genome-wide and suggestive AD-related genetic associations extracted from GWAS publications. This includes 6,990 genetic associations for variants, genes, and SNP interactions. Figure 2 shows the distribution of ADVP genetic associations by harmonized meta-information data fields: a) Nine harmonized phenotypes; b) Six harmonized analyses type; c) Population, and d) Cohorts/Consortiums.

**Figure 2:**
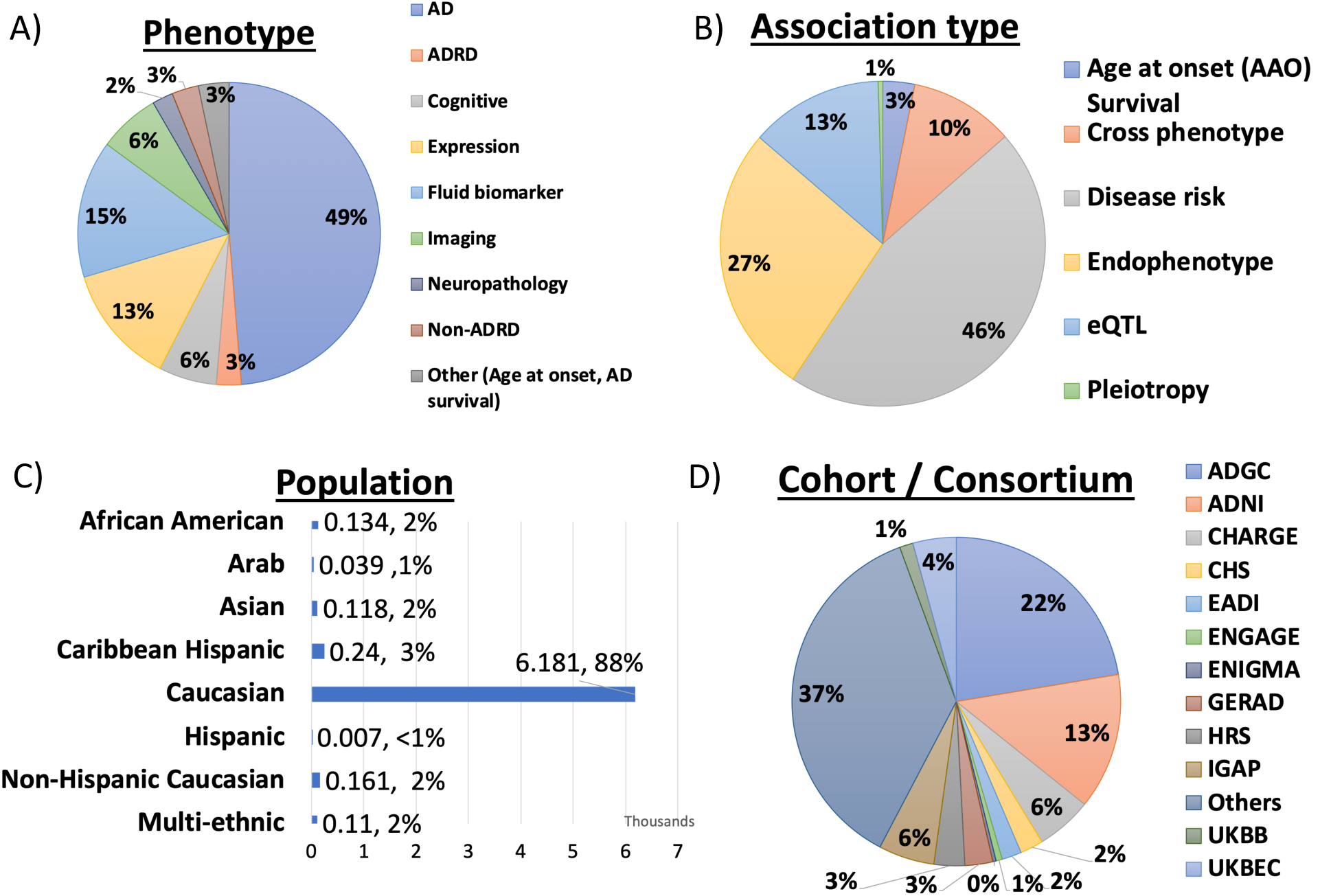
Summary of genetic association records in ADVP by **A**) Phenotype, **B**) Association type, **C**) Population, and **D**) Cohort/Consortium.

All ADVP association records are uniquely standardized into different categories:

1. As shown in Figure 2A, ADVP records are associated with nine different phenotype categories, with roughly half of them related to AD diagnosis. 15% of the records are related to fluid biomarkers, 7% with imaging and 6% with cognitive measures.
2. With respect to analysis type categories, ADVP includes 3,199 (45.8%) association records reported in disease-risk analyses, of which 1,342 and 934 associations are reported by meta- and joint-analyses, respectively. 1,887 (26.9%) of the records are related to AD endophenotype and 924 (13.2%) eQTL AD associations (Figure 2B).
3. ADVP is the first to collect AD genetic associations at SNP level (6,437, 92.1%), gene level (320, 4.5%), as well as SNP and gene interactions (233, 3.3%).
4. ADVP records present analyses results from seven populations as well as those from multi-ethnic analyses. ∼88% of the records are for Caucasian (Figure 2C). Others include African American, Arab, Asian, Caribbean Hispanic, Hispanic and Non-Hispanic Caucasian.
5. ADVP records span analyses results published by ADGC using over 80 cohorts (Figure 2D), including ADGC, Alzheimer’s Disease Neuroimaging Initiative (ADNI), Cohorts for Heart and Aging Research in Genomic Epidemiology (CHARGE) Consortium, European Association of Development Research and Training Institutes (EADI), European Network for Genetic and Genomic Epidemiology (ENGAGE) Consortium, Enhancing NeuroImaging Genetics through Meta-Analysis (ENIGMA) Consortium, The International Genomics of Alzheimer’s Project (IGAP) and others. See **Supplementary Table S3** for details on cohorts included in ADVP.

Furthermore, ADVP provides annotation information for each genetic association (Section “**2.4 Annotation**”). In summary, all the genetic association records in ADVP were represented by >1,800 unique variants (based on genomic position) and >900 genomic loci (based on computed normalization). ADVP associations are mostly located in non-coding regions including intronic (52.9%), intergenic (15.2%), and promoter (5.9%) (Figure 3A). ADVP records are also cross-referenced to NIAGADS Alzheimer’s Genomics database [24]. Figure 3B summaries genetic variants in ADVP by functional impact as determined by ADSP functional annotation pipeline [24,37,38] that uses VEP effect predictor with customized ranking to generate most damaging consequence for each variant [38].

**Figure 3:**
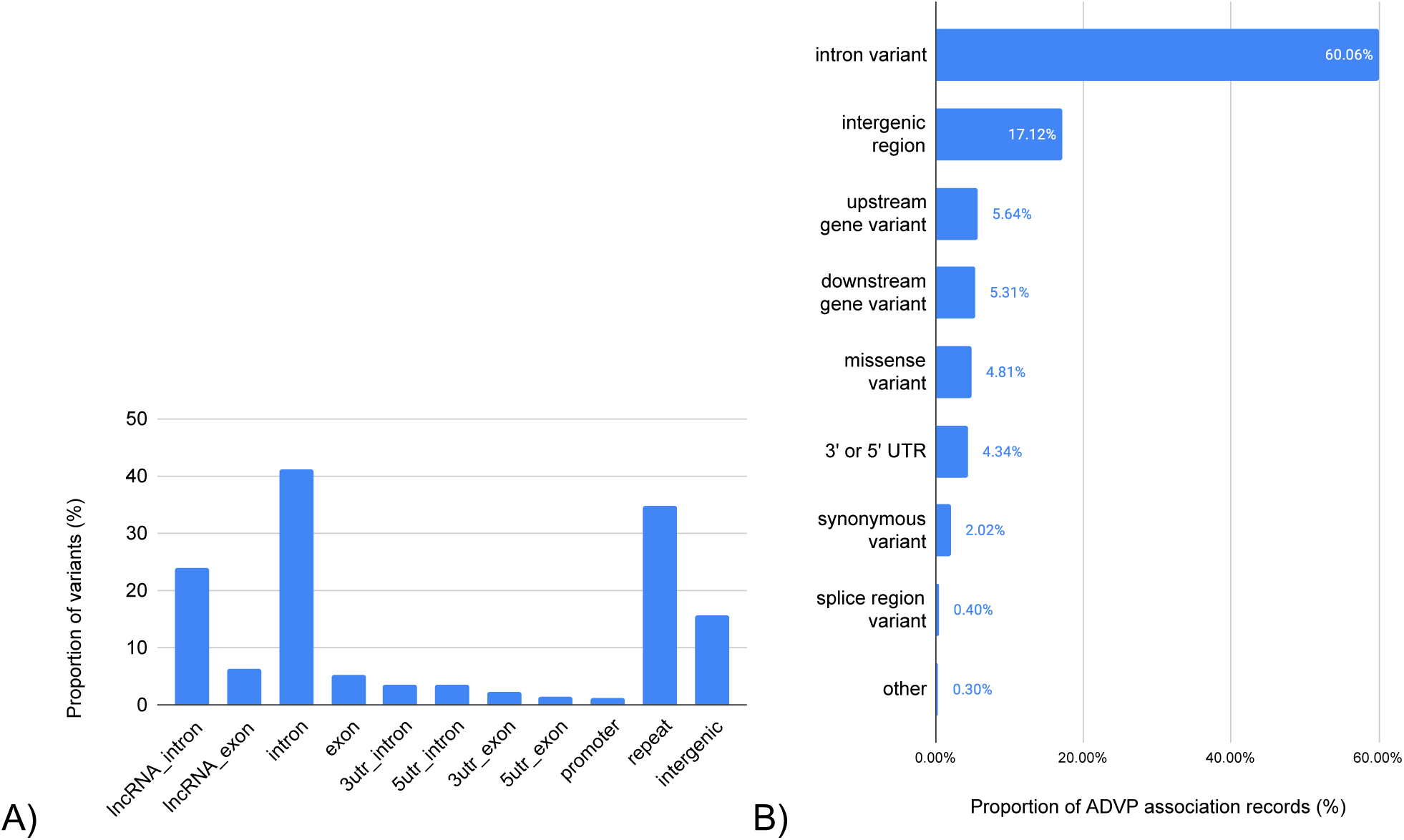
Summary of ADVP association records by genome annotation and most-significant functional consequence. **A)** Genomic localization of ADVP variants within mRNA, lncRNA, and repeat elements. Shown are proportion (%) of variants in each genomic element category; **B)** Most-significant predicted variant impact. Impact for variants is determined using ADSP functional annotation pipeline [38, 57] and is provided by NIAGADS Alzheimer’s Genomics database [24]. The consequence for a variant is predicted and ranked based on multiple criteria including genomic location of a variant, genes, transcripts and protein sequences, biological type of transcript, transcript support level and other factors.

### 3.2 ADVP features – search, browse and visualize

ADVP aims to provide a simple and unified resource to the scientific community, allowing investigators to search and browse AD genetic association information more easily. This is first done by displaying association records using a pre-selected set of most important data fields (Section “**2.3.2.2 Description of association records**”). Investigators can further select additional data fields via the column selector (Figure 4A). All records are integrated with the NIAGADS Alzheimer’s Genomics database, allowing investigators to explore various kinds of biological annotations (e.g., CADD score (Rentzsch et al., 2019)) and functional genomics evidence, including overlaps with FANTOM5 [35], ENCODE histone modification [40], and gene ontologies from KEGG [41] and UniProt [42].

**Figure 4.**
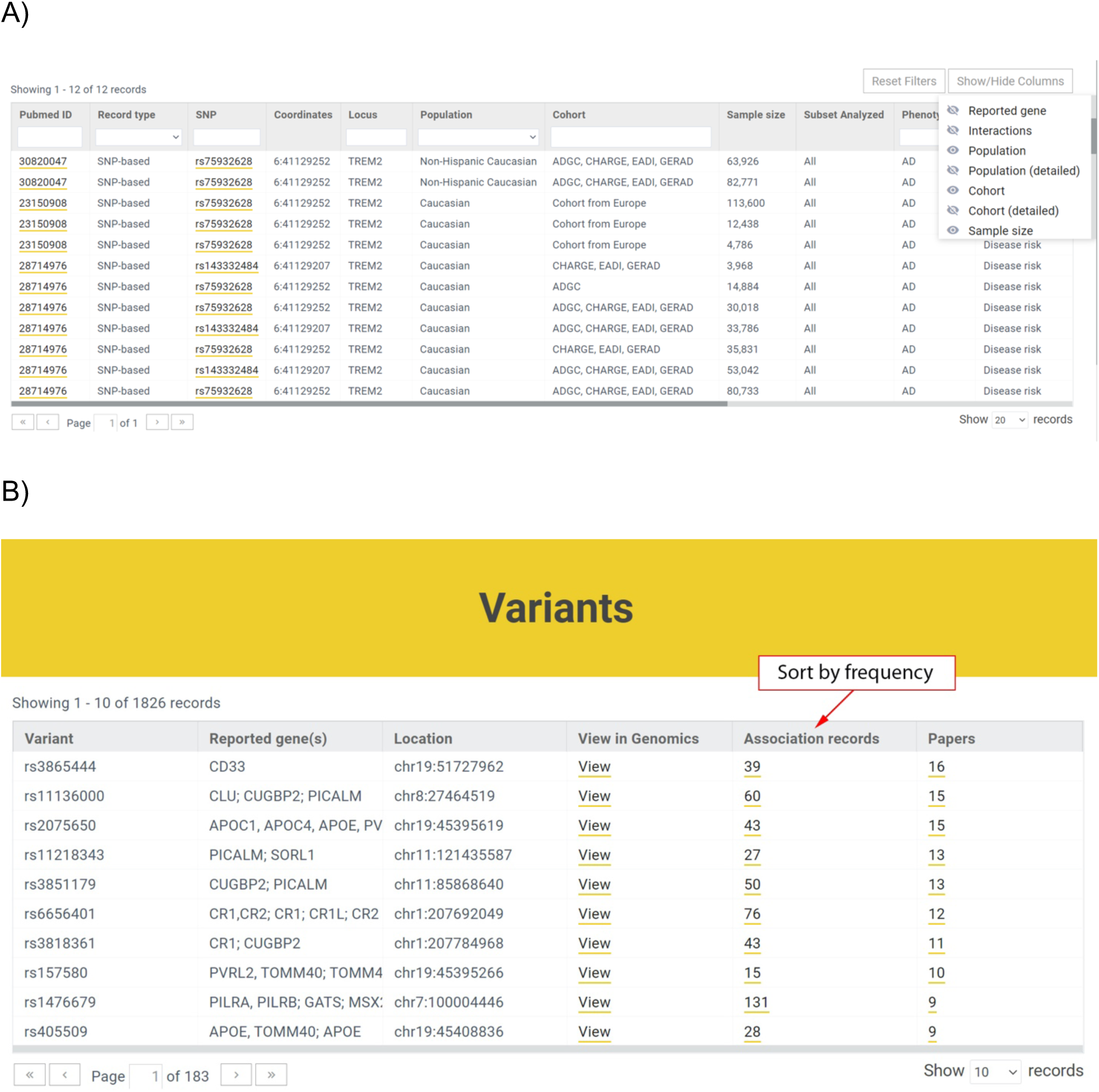
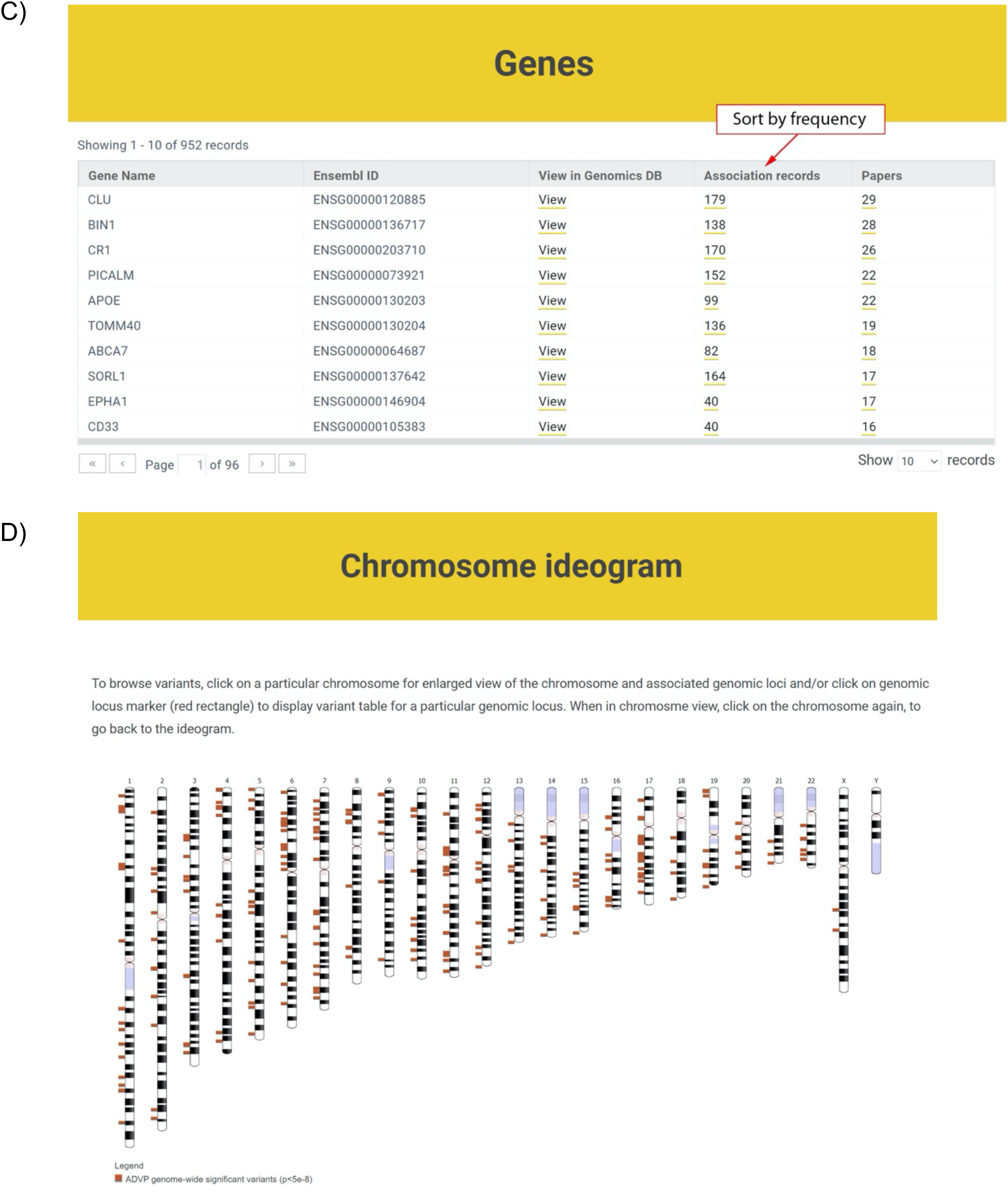
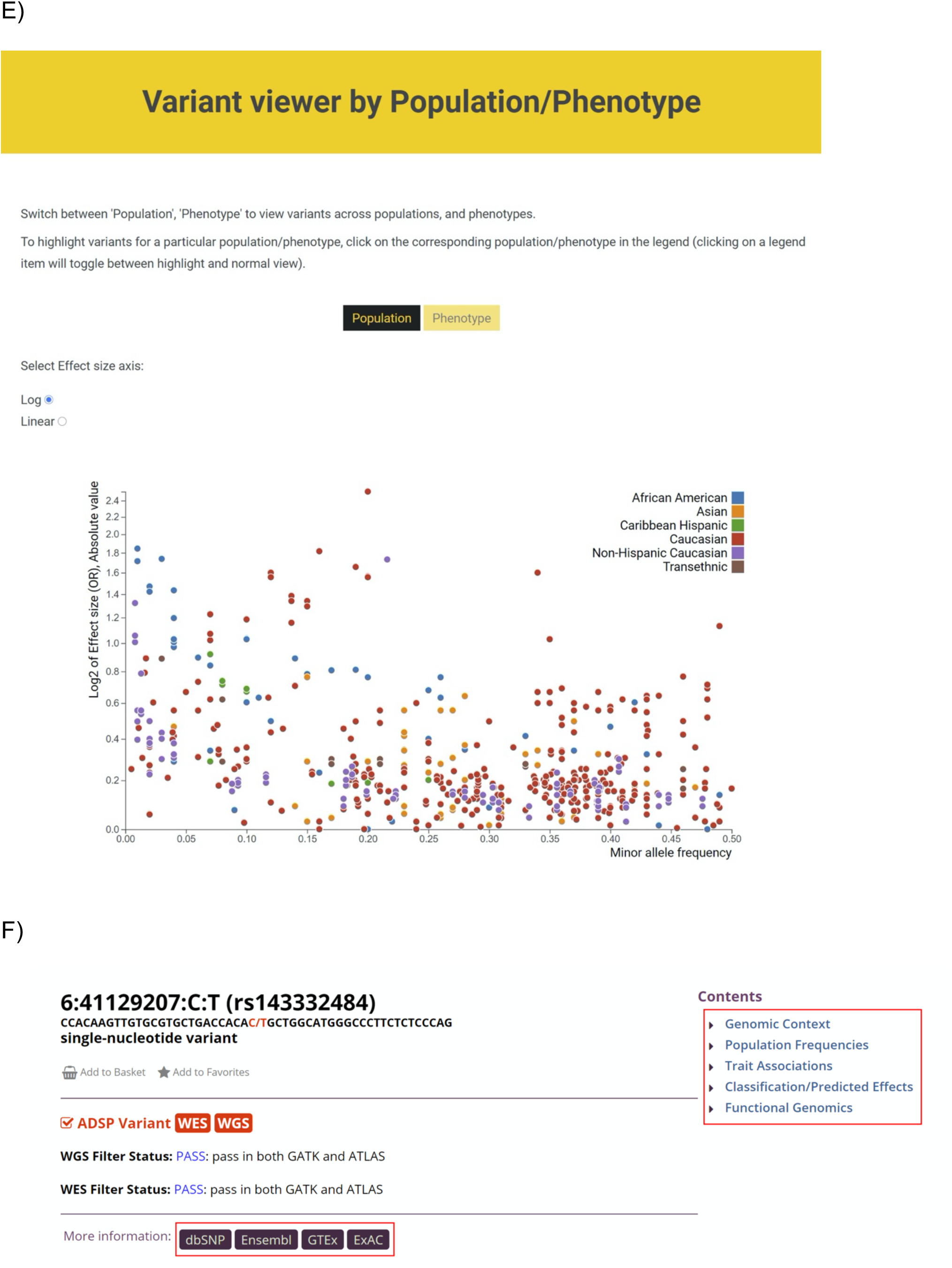
ADVP interface. **A)** Association records table. The displayed information can be customized via column/field selector and filtered using provided text and data filters; **B)** Top variants curated in ADVP. Variants are displayed according to the number of reporting publications by default; association records for variants and variant-related publications can be quickly accessed; **C)** Top genes curated in ADVP. Genes are ordered by the number of reporting publications by default; gene-related association records and publications can be quickly accessed; **D)** Interactive chromosome ideogram-based view of association data; **E)** Interactive variant viewer by population and phenotype. Variants are arranged by their effect size (odds ratio; Y-axis) and allele frequency (X-axis) and color-coded by population and phenotype; **F)** Integration with NIAGADS AD Genomics database [24] providing additional biological information and functional evidence (red rectangles).

The ADVP search interface was designed based on focus group use cases. ADVP provides several ways to search for genetic association records:

1. By publication – investigators can quickly identify and retrieve all association records curated by ADVP for a particular study using PMID, PMCID, first or last author names, year of publication or article title (https://advp.niagads.org/publications).
2. By variant or gene of interest – investigators can search for the variant (https://advp.niagads.org/variants) or gene (https://advp.niagads.org/genes) of interest and browse all the associated ADVP records. Additionally, investigators can easily discover top variants or a gene with most association records or most publications via the summary counts for association records and papers (Figure 4B, 4C).
3. By region of interest – investigators can search and retrieve all genetic associations within the genomic regions of interest (https://advp.niagads.org/search).
4. By integrative genome-wide plots – investigators can navigate the landscape of AD genetics associations using the interactive chromosome ideogram (https://advp.niagads.org/ideogram, Figure 4D) or interactive population/phenotype variant viewer (https://advp.niagads.org/plot, Figure 4E).

Additionally, for each variant or gene, users can view the biological annotations and functional evidence (e.g., 1000 Genomes project [31], dnSNP [25], GTEx [32], ENCODE [40] and others) via the NIAGADS Alzheimer’s Genomics database [24] (Figure 4F).

### 3.3 ADVP use cases

ADVP is designed for various use cases in mind. First, ADVP can serve as a point of entry for investigators to explore the AD genetics literature. They can browse through variant and gene records, identify top associated loci for particular populations and phenotypes, or inspect top GWAS associations in the gene or genomic region of interest. Second, investigators can use ADVP to check their association analysis findings. They can further restrict the comparison by focusing on results from a specific population, cohort, or by comparing the strength of associations via *p*-values or phenotypes. Lastly, investigators can use ADVP to check if their findings have functional support from eQTL or other functional evidence collected in the NIAGADS Alzheimer’s genomics database (Figure 4F).

### 3.4. Genetic architecture of AD-associated loci across populations

To show the diversity and breadth of ADVP data, we performed population-based analysis of AD associations in ADVP (Section **“2.6 Population-based analysis of AD associations”**). Across four major populations (African American, Asian, Caribbean Hispanic, Caucasian/Non-Hispanic white) with the most association records, 91 loci (Section “**2.6 Population-based analysis of AD associations”**) were identified in any of these 4 populations, whereas 10 of them were found in two or more populations, including *BIN1*, *CD33*, *PICALM*, *SORL1*, and *ABCA7*. The majority of AD loci (81/91=89%) were population-specific, i.e. found in only one population. This could partially be explained by the underlying genetic differences across populations, but could also be contributed by variability in GWAS sample sizes across studies/populations, which could lead to the observed differences in association strength and heterogeneity of loci identified in each population. Across the four populations, the corresponding top SNP (Section “**2.6 Population-based analysis of AD associations”**), effect size and allele frequency on all these 91 loci are available in **Supplementary Table S4**.

We next explored the effect size and allele frequency of all AD-associated loci that are found in any populations (Figure 5A), or those that are found in two or more populations (shared AD loci) (Figure 5B).

**Figure 5.**
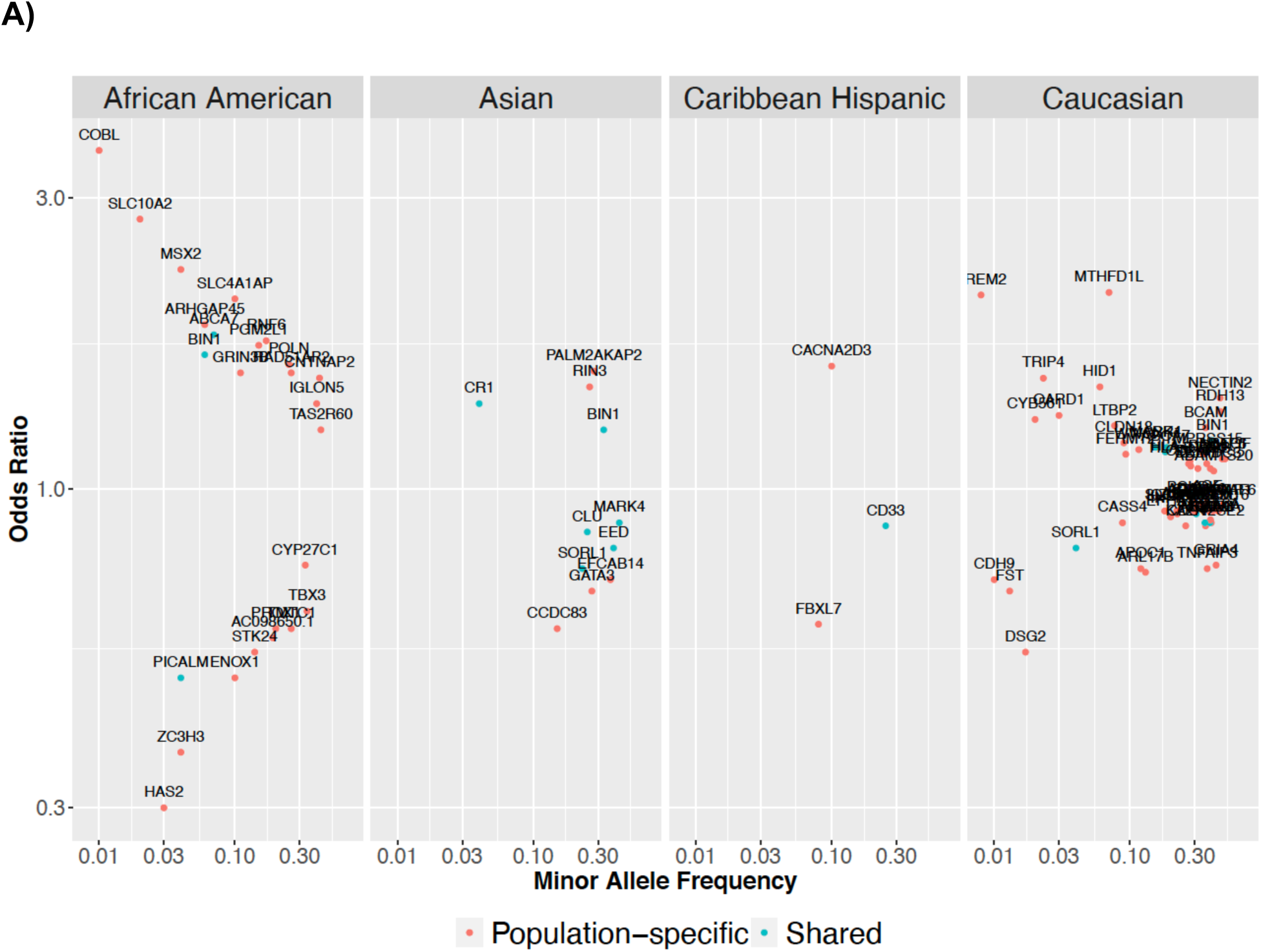
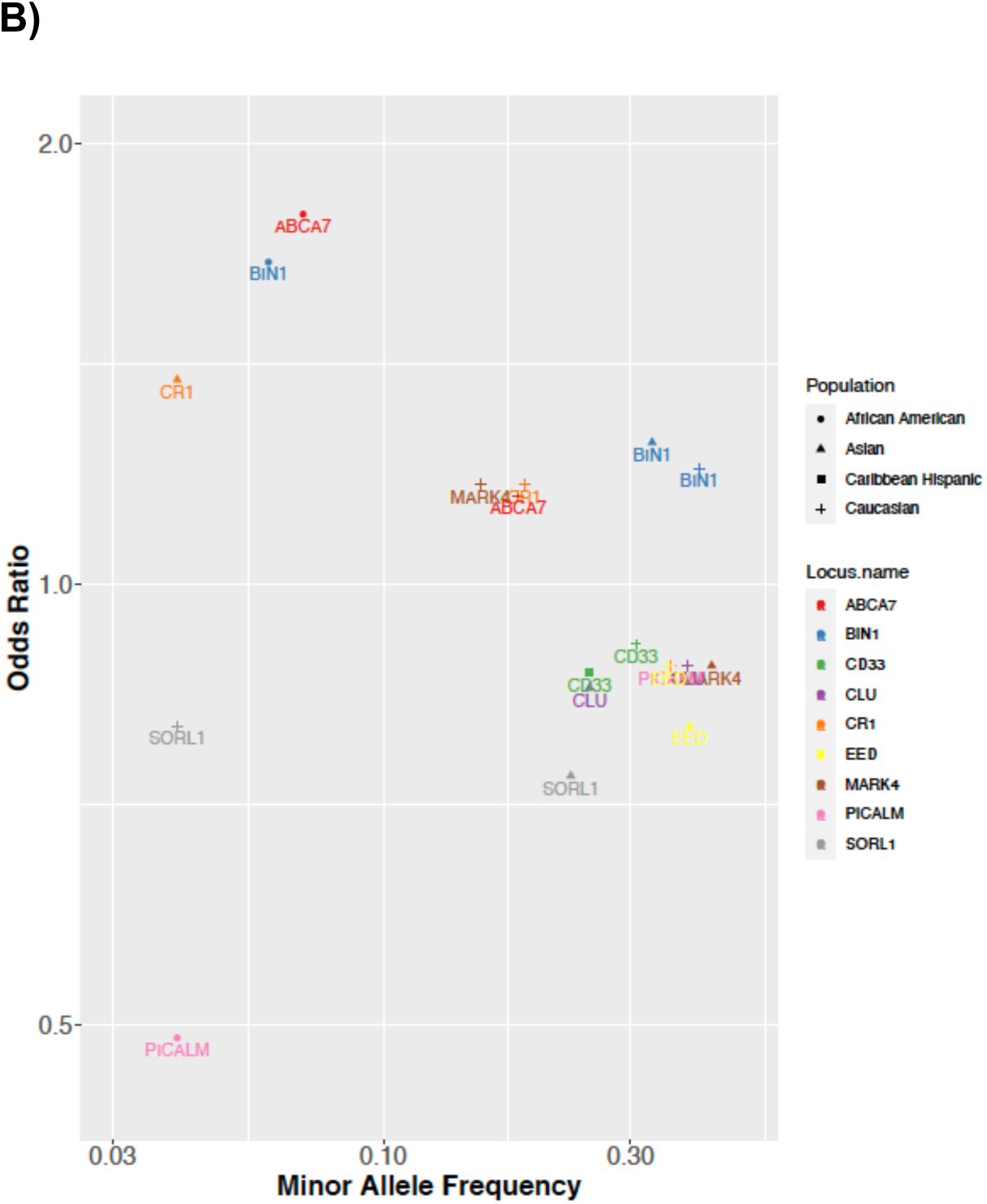
ADVP catalogs AD genetic associations across 8 populations (>80 cohorts). AD-associated loci (non-*APOE*) for four major populations (African American, Asian, Caribbean Hispanic, Caucasian/Non-Hispanic White) are shown in this figure. For each locus, shown are the allele frequency and odds ratio for the most significant variant located with the locus. ADVP data captures the diversity and population-specificity of AD-associated loci. **A)** Shown are loci associated with AD in each of the major populations (shared loci are colored black, population-specific loci are shown in blue). **B)** Shown are shared AD loci reported in two or more major populations.

As can be seen from Figure 5, ADVP captures reported AD-associated loci across many populations (Figure 5A; only non-*APOE* loci from 4 populations are shown). ADVP contains both population-specific AD loci and loci that are shared across populations. As shown in Figure 5B, the shared loci tend to preserve their risk (e.g., *ABCA7* [43]) and protective roles (e.g., *PICALM* [44]).

On the other hand, as shown in Figure 5A, common loci tend to exert smaller effects compared to population-specific loci, yet these effects of the common loci vary across populations (Figure 5B) (i.e. are population-specific).

### 3.5. Functional analysis of ADVP variants

We next investigated the functional impact of ADVP variants. To do so, we analyzed all ADVP variants that are associated with AD/ADRD (excluding eQTLs) in the Caucasian or non-Hispanic White populations. A total of 1,675 ADVP variants met these criteria and were analyzed.

First, to determine the potential causal genes for these variants, we asked if these variants regulate any genes in any AMP-AD [45] eQTL data [46] obtained from three different brain regions (dorsolateral prefrontal cortex, cerebellum and temporal cortex). Please see this publication [46] for details of how these data eQTL was harmonized and processed.

31% of the analyzed ADVP variants were identified as significant eQTLs in at least one of the three AMP-AD eQTL datasets (FDR<0.01), and 32% among these were eQTLs in all three brain regions. Altogether, these variants targeted 130 genes (including HLA region), of which 31 were also the nearest genes reported in ADVP. These target genes (e.g., *ACE, PVR*) were enriched in cell junction organization [47], and acetyltransferase activity (cholinergic system) [48], which recapitulated some of the molecular and cellular mechanisms underlying the pathogenesis of AD.

Second, we analyzed and ranked relevant tissue categories implicated by ADVP variants. To do so, we quantified the significance and enrichment of tissue-specific enhancer overlaps with ADVP variants (Section “**2.7 Functional analysis of AD associations”**). The ADVP variants were enriched in regulatory elements profiled by FANTOM5 and Roadmap Epigenomics (ChromHMM). Top tissues enriched in overlaps between ADVP variants and Roadmap enhancers included AD-relevant blood (OR=3.7), digestive (OR=3.5), brain (OR=2.8), and skeletal (OR=2.6) tissue categories. The overlaps between ADVP variants and enhancers were shown to be enriched in tissues known to be related to AD in various ways [49–55]. For instance, the implicated immunity-related blood category is in line with recent work highlighting the role of neuroinflammation in AD pathogenesis and etiology [49, 55]. Digestive is related to the gut microbiome, which can be linked to AD behavior in mice [56]. The implicated skeletal category has also been previously linked with brain atrophy in AD [51].

In summary, analyses of genetic associations in ADVP revealed potential functional roles of the AD variants in relevant tissue/cell type context, and recapitulated some of the known regulatory mechanisms underlying AD pathogenesis.

## 4. Discussion

Here, we present ADVP, a portal to search, browse and visualize the largest collection of systematically curated, harmonized, and annotated AD-specific genetic variants and associations (∼7,000 genetic associations in the current V1.0 release, November 2020). Among the main distinctive features of ADVP is the uniqueness of reporting harmonized AD variant and association information (standardized meta-table curation schema), integration with the genomic annotation, and functional information (NIAGADS Alzheimer’s Genomics database [24]), as well as extensive cohort/consortium level information.

ADVP uniquely includes associations at SNP, gene, and interaction levels and contains curated phenotypes not limited to disease risk, but also includes endophenotypes, fluid biomarkers, imaging, neuropathology, and other phenotypes. Moreover, ADVP curates and records AD and ADRD eQTL association findings (Figure 2B).

In addition to the standard *p*-values and effect sizes reported for association records, ADVP puts particular emphasis on harmonizing meta-data curated from the publications. Both the curated and derived columns are stored in the database. These include phenotype, association type, standardized gene names, study information (population, cohort, sample size, subset analyzed), and details of analyses (analyses type, imputation) (Figure 4A). All these columns enable the investigators to interpret, compare and view these records at different levels: phenotype (Figure 2A), population (Figure 2C), cohort (Figure 2D), to name a few.

All ADVP records are annotated with the genomic context (upstream/downstream genes, and their distances) and their co-localized genomic element (Figure 3). They are also cross-referenced with NIAGADS Alzheimer’s Genomics DB [24], providing other genomic annotation and functional genomic information. The standardized, structured design of ADVP association data allows systematic integration with other genetic, genomics, and molecular databases.

Population-based and functional analyses of AD associations revealed the genetic architecture of AD-associated loci and points to tissue-specific regulatory mechanisms for AD. However, variability in GWAS sample sizes may contribute to the observed differences in associations and loci across different populations. Broadening the ADVP coverage of population groups, as well as expanding functional data types and coverage will provide further insights in complex genetic architecture and biology underlying AD.

Lastly, we made substantial efforts to ensure high-quality of ADVP data contents. First, quality control at multiple levels is performed (Figure 1, Section “**2.5 Quality control steps**”) to ensure the uniqueness of included genetic associations (no double counting / re-reporting of associations). Besides, variant information in ADVP has been cross-checked against other reference databases such as dbSNP.

ADVP will continuously be updated with versioned releases every six months. New publications on AD-related GWASs and corresponding associations will be added in an ongoing manner.

In the future, ADVP data collections will consist of a broader range of genetic results:

1. AD whole-genome/whole-exome sequencing analyses
2. AD xQTL associations, where x = protein, methylation, epigenetics marks, or other molecular traits Other genetic variant types, such as insertions/deletions (indel), copy number variations (CNV), or structural variations (SV) as they become available
3. AD related dementias (ADRD) and neurodegenerative disorders

Future ADVP functionality will include further collection and addition of functional genomic evidence supporting genetic associations.

To conclude, ADVP contains the largest collection of systematically curated, harmonized, and annotated literature-derived variants for AD to the best of our knowledge. The extensive and unique features in ADVP allow investigators to easily access, interpret, compare, and visualize the vast collection of AD genetics findings.

## 5. Availability

All AD variant and association information is available through ADVP website (https://advp.niagads.org). The code for processing reported variant and association data is available upon request.

## Supporting information

All supplementary information

## Data Availability

Data is available at https://advp.niagads.org.

## Acknowledgement

The authors thank Laura Cantwell, Lauren Kleidermacher, and Mitchell Tang for their contributions at various stages of this work. This work was supported by the National Institute on Aging [U24-AG041689, U54-AG052427, U01-AG032984, U01AG058654]; Biomarkers Across Neurodegenerative Diseases (BAND 3) (award number 18062), co-funded by Michael J Fox Foundation, Alzheimer’s Association, Alzheimer’s Research UK and the Weston Brain institute.

