## Supplementary material for "Alzheimer’s Disease variant portal (ADVP): a catalog of genetic findings for Alzheimer’s Disease": All supplementary information

**Supplementary Methods Note 1** – Derivation and full description of the nine meta-information data fields describing each association record
**Supplementary Methods Note 2** – ADVP front-end and back-end architecture and implementation
**Supplementary Table 1** – Curated publications and data sources
**Supplementary Table 2** – All curated and harmonized/derived data fields in ADVP
**Supplementary Table 3** – ADGC cohorts included in ADVP
**Supplementary Table 4** – Top AD-associated loci and SNPs across populations
**Supplementary Figure 1** – Classification of studies by the analysis type and stage of analyses

### Supplementary Methods

#### Note 1

Details on the nine meta-information data fields describing each association:

1) "Record type": association record type is set based on whether the reported association is for a single SNP ("SNP-level"), a single gene ("Gene-level"), SNP interactions ("Interaction (SNP)", or gene interactions ("Interaction (Gene)").
2) "Population": study population information was first copied from the publication ("Population (detailed)"). Then, the reported population information was further mapped using standard population vocabulary to normalize population information across studies ("Population" column).
3) "Cohort" - Full cohort names ("Cohort (detailed)") were mapped to consortium names whenever available to obtain normalized cohort information ("Cohort"). For example, if the cohort is one of the ADGC GWAS cohorts (see **eTable 3**), ADVP appends "ADGC" to the data entry.
4) "Sample size": Original sample size (number of cases, number of controls when available/applicable) were recorded. ADVP V1.0 webserver only reports the total number.
5) "Subset analyzed": description of the subset of samples used in association analysis. When a subset of samples was used to perform association analysis (e.g., "e2/e4", "e4 carriers only", "Female only"), this field records description of the subset as described in the publication.
6) "Phenotype": the outcome variable (i.e., phenotype/trait) of the association analysis. Original outcome ("Phenotype (detailed)") was assigned to one of the nine categories including "AD", ADRD", "Cognitive", "Expression", "Fluid biomarker", "Imaging", "Neuropathology", Non-ADRD", and "Other" (representing "Age of onset" or "AD survival").
7) "Association Type": the type of association analysis. Based on the "Phenotype" column, we classified each association test into six categories: "Age at onset (AAO)/ Survival", "Cross phenotype", "Disease-risk", "Endophenotype", expression quantitative trait locus "eQTL", "Pleiotropy".
8) "Stage": the stage of the analysis as described in the publication, e.g., "Stage n" (n=1,2,3), "Discovery", "Validation", "Meta-analysis". The stage information is reported as given in the paper if this information was available. If the stage information was not explicitly provided in the text, we derived the stage information as follows (**eFigure 1**). If the association analysis was done using a single cohort, the "Stage" of the record was set as: "Stage n" (n=1,2,3 or others); "Discovery", "Replication/Validation". "Discovery" was used if the paper was the first to report such findings using the specific combination of cohort + phenotype information, otherwise, the stage was set as "Replication/Validation". If the association analysis was done using multiple cohorts, the stage was set as "Meta-analysis" if the analyses were performed using methods such as "inverse-variance weighting", "fixed effects", "random effects" model or METAL R package; if not, the stage was set to "Joint-analysis".
9) "Imputation": imputation panel information. The imputation panel version, software tool and version were mapped to broader categories such as 1000 Genome project ("1000G"), International HapMap Project ("HapMap") or Haplotype Reference Consortium ("HRC").

#### Note 2

**ADVP front-end and back-end architecture and implementation**

ADVP is designed with ease of update and modularity in mind. Contents of ADVP are derived from collection, curation, harmonization, processing and integration of AD-related publications and reported genetic associations using a standardized meta-data schema (see **Section 2.3 Meta-data curation** in the main text). The ADVP web server runs on Amazon Web Services (AWS) cloud computing instance (m5.4xlarge) using MySQL [33] relational database management system as a back-end and a PHP/JQuery-based web front-end. All the publication, variant and association information stored in ADVP relational database is organized into multiple tables (**Figure 1** in the main text) including publications, variant and association tables. The web front-end provides multiple data views for publications, genes, variants, and association records (**Figure 4** in the main text).

**Supplementary Table 1 – Curated publications and data sources**

| Pubmed PMID | Year | First author | Last author | Title | Journal | Vol/Issue/Page | Curated tables | Curation source |
|---|---|---|---|---|---|---|---|---|
| 20697030 | 2010 | Jun G | Schellenberg GD | Meta-analysis confirms CR1, CLU, and PICALM as alzheimer disease risk loci and reveals interactions with APOE genotypes. | Archives of neurology | 67(12):1473-84 | Table 3,5 | ADGC |
| 20879451 | 2010 | Shen L | Saykin AJ | Sparse bayesian learning for identifying imaging biomarkers in AD prediction. | Medical image computing and computer-assisted intervention : MICCAI ... International Conference on Medical Image Computing and Computer-Assisted Intervention | 13(Pt 3):611-8 | NR | ADGC |
| 21460841 | 2011 | Naj AC | Schellenberg GD | Common variants at MS4A4/MS4A6E, CD2AP, CD33 and EPHA1 are associated with late-onset Alzheimer's disease. | Nature genetics | 43(5):436-41 | Table 1,2 | ADGC |
| 23565137 | 2013 | Miyashita A | Kuwano R | SORL1 is genetically associated with late-onset Alzheimer's disease in Japanese, Koreans and Caucasians. | PloS one | 8(4):e58618 | Table 2,3,4 | ADGC |
| 23571587 | 2013 | Reitz C | Mayeux R | Variants in the ATP-binding cassette transporter (ABCA7), apolipoprotein E 4,and the risk of late-onset Alzheimer disease in African Americans. | JAMA | 309(14):1483-92 | Table 2,3 | ADGC |
| 23673467 | 2013 | Reitz C | Mayeux R | Independent and epistatic effects of variants in VPS10-d receptors on Alzheimer disease risk and processing of the amyloid precursor protein (APP). | Translational psychiatry | 3:e256 | Table 2 | ADGC |
| 23700308 | 2013 | Leung YY | Wang LS | CoRAL: predicting non-coding RNAs from small RNA-sequencing data. | Nucleic acids research | 41(14):e137 | NR | ADGC |
| 23727082 | 2014 | Carney RM | Pericak-Vance MA | Parkinsonism and distinct dementia patterns in a family with the MAPT R406W mutation. | Alzheimer's & dementia : the journal of the Alzheimer's Association | 10(3):360-5 | NR | ADGC |
| 23759147 | 2013 | Barral S | Mayeux R | Exceptional memory performance in the Long Life Family Study. | Neurobiology of aging | 34(11):2445-8 | NR | ADGC |
| 23836404 | 2013 | Shulman JM | Bennett DA | Genetic susceptibility for Alzheimer disease neuritic plaque pathology. | JAMA neurology | 70(9):1150-7 | Table 1,2,3 | ADGC |
| 23845100 | 2013 | Nuytemans K | Vance JM | C9ORF72 intermediate repeat copies are a significant risk factor for Parkinson disease. | Annals of human genetics | 77(5):351-63 | NR | ADGC |
| 23894628 | 2013 | Kim S | Saykin AJ | Influence of genetic variation on plasma protein levels in older adults using a multi-analyte panel. | PloS one | 8(7):e70269 | Table 3,4 | ADGC |
| 23943636 | 2013 | Lin CF | Wang LS | DRAW+SneakPeek: analysis workflow and quality metric management for DNA-seq experiments. | Bioinformatics (Oxford, England) | 29(19):2498-500 | NR | ADGC |
| 24086677 | 2013 | Honea RA | Goate AM | Characterizing the role of brain derived neurotrophic factor genetic variation in Alzheimer's disease neurodegeneration. | PloS one | 8(9):e76001 | Table 2,3 | ADGC |
| 24098339 | 2013 | Cao K | Wang LS | Analysis of nonlinear gene expression progression reveals extensive pathway and age-specific transitions in aging human brains. | PloS one | 8(10):e74578 | NR | ADGC |
| 24131184 | 2013 | Reitz C | Mayeux R | TREM2 and neurodegenerative disease. | The New England journal of medicine | 369(16):1564-5 | NR | ADGC |
| 24145223 | 2014 | Ryvkin P | Wang LS | Using machine learning and high-throughput RNA sequencing to classify the precursors of small non-coding RNAs. | Methods (San Diego, Calif.) | 67(1):28-35 | NR | ADGC |
| 24162737 | 2013 | Lambert JC | Amouyel P | Meta-analysis of 74,046 individuals identifies 11 new susceptibility loci for Alzheimer's disease. | Nature genetics | 45(12):1452-8 | Table 2 | ADGC |
| 24209629 | 2013 | Guerreiro R | Hardy J | SnapShot: genetics of Alzheimer's disease. | Cell | 155(4):968-968.e1 | NR | ADGC |

| Pubmed PMID | Year | First author | Last author | Title | Journal | Vol/Issue/Page | Curated tables | Curation source |
|---|---|---|---|---|---|---|---|---|
| 24244562 | 2013 | Ridge PG | Kauwe JS | Alzheimer's disease: analyzing the missing heritability. | PloS one | 8(11):e79771 | Table 1 | ADGC |
| 24439028 | 2014 | Logue MW | Farrer LA | Search for age-related macular degeneration risk variants in Alzheimer disease genes and pathways. | Neurobiology of aging | 35(6):1510.e7-18 | Table 1,3,4,5 | ADGC |
| 24439484 | 2014 | Benitez BA | Cruchaga C | Missense variant in TREML2 protects against Alzheimer's disease. | Neurobiology of aging | 35(6):1510.e19-26 | Table 1 | ADGC |
| 24495969 | 2014 | Ruiz A | Ramirez A | Follow-up of loci from the International Genomics of Alzheimer's Disease Project identifies TRIP4 as a novel susceptibility gene. | Translational psychiatry | 4:e358 | Table 1 | ADGC |
| 24510649 | 2013 | Lin CF | Wang LS | Analyzing copy number variation using SNP array data: protocols for calling CNV and association tests. | Current protocols in human genetics | 79:Unit 1.27. | NR | ADGC |
| 24755620 | 2014 | Perez-Palma E | De Ferrari GV | Overrepresentation of glutamate signaling in Alzheimer's disease: network-based pathway enrichment using meta-analysis of genome-wide association studies. | PloS one | 9(4):e95413 | Table 2 | ADGC |
| 24770881 | 2014 | Nelson PT | Fardo DW | ABCC9 gene polymorphism is associated with hippocampal sclerosis of aging pathology. | Acta neuropathologica | 127(6):825-43 | Table 3,4 | ADGC |
| 25419706 | 2014 | Wetzel-Smith MK | Graham RR | A rare mutation in UNC5C predisposes to late-onset Alzheimer's disease and increases neuronal cell death. | Nature medicine | 20(12):1452-7 | NR | ADGC |
| 25470345 | 2015 | Nelson PT | Fardo DW | Reassessment of risk genotypes (GRN, TMEM106B, and ABCC9 variants) associated with hippocampal sclerosis of aging pathology. | Journal of neuropathology and experimental neurology | 74(1):75-84 | Table 2 | ADGC |
| 25531812 | 2015 | Wang LS | Yu L | Rarity of the Alzheimer disease-protective APP A673T variant in the United States. | JAMA neurology | 72(2):209-16 | NR | ADGC |
| 25533204 | 2015 | N | A | Convergent genetic and expression data implicate immunity in Alzheimer's disease. | Alzheimer's & dementia : the journal of the Alzheimer's Association | 11(6):658-71 | NR | ADGC |
| 25649651 | 2015 | Wang X | Kamboh MI | Genetic Determinants of Survival in Patientswith Alzheimer's Disease. | Journal of Alzheimer's disease : JAD | 45(2):651-8 | Table 3,4 | ADGC |
| 25663231 | 2015 | Beecham GW | Vance JM | PARK10 is a major locus for sporadic neuropathologically confirmed Parkinson disease. | Neurology | 84(10):972-80 | Table 2 | ADGC |
| 25687773 | 2015 | Desikan RS | Dale AM | Genetic overlap between Alzheimer's disease and Parkinson's disease at the MAPT locus. | Molecular psychiatry | 20(12):1588-95 | Table 2 | ADGC |
| 25762156 | 2015 | Malik M | Estus S | Genetics of CD33 in Alzheimer's disease and acute myeloid leukemia. | Human molecular genetics | 24(12):3557-70 | Table 1 | ADGC |
| 25778476 | 2016 | Jun G | Farrer LA | A novel Alzheimer disease locus located near the gene encoding tau protein. | Molecular psychiatry | 21(1):108-17 | Table 1,2 | ADGC |
| 25862742 | 2015 | Desikan RS | Dale AM | Polygenic Overlap Between C-Reactive Protein, Plasma Lipids, and Alzheimer Disease. | Circulation | 131(23):2061-2069 | Table 2,3 | ADGC |
| 26049409 | 2015 | Hirano A | Kanba S | A genome-wide association study of late-onset Alzheimer's disease in a Japanese population. | Psychiatric genetics | 25(4):139-46 | Table 2,3 | ADGC |
| 26079416 | 2015 | Mukherjee S | Glymour MM | Genetically predicted body mass index and Alzheimer's disease-related phenotypes in three large samples: Mendelian randomization analyses. | Alzheimer's & dementia : the journal of the Alzheimer's Association | 11(12):1439-1451 | Table 4 | ADGC |
| 26079503 | 2015 | Ostergaard SD | Scott RA | Associations between Potentially Modifiable Risk Factors and Alzheimer Disease: A Mendelian Randomization Study. | PLoS medicine | 12(6):e1001841; discussion e1001841 | NR | ADGC |
| 26092349 | 2016 | Hohman TJ | Thornton-Wells TA | Global and local ancestry in African-Americans: Implications for Alzheimer's disease risk. | Alzheimer's & dementia : the journal of the Alzheimer's Association | 12(3):233-43 | Table 2 | ADGC |

| Pubmed PMID | Year | First author | Last author | Title | Journal | Vol/Issue/Page | Curated tables | Curation source |
|---|---|---|---|---|---|---|---|---|
| 26339675 | 2015 | Tosto G | Mayeux R | F-box/LRR-repeat protein 7 is genetically associated with Alzheimer's disease. | Annals of clinical and translational neurology | 2(8):810-20 | Table 2,3 | ADGC |
| 26365416 | 2016 | Kunkle BW | Pericak-Vance MA | Genome-wide linkage analyses of non-Hispanic white families identify novel loci for familial late-onset Alzheimer's disease. | Alzheimer's & dementia : the journal of the Alzheimer's Association | 12(1):2-10 | Table 2 | ADGC |
| 26366463 | 2015 | Ghani M | Rogaeva E | Association of Long Runs of Homozygosity With Alzheimer Disease Among African American Individuals. | JAMA neurology | 72(11):1313-23 | NR | ADGC |
| 26433351 | 2015 | Barral S | Mayeux R | Linkage analyses in Caribbean Hispanic families identify novel loci associated with familial late-onset Alzheimer's disease. | Alzheimer's & dementia : the journal of the Alzheimer's Association | 11(12):1397-1406 | Table 2,3,4 | ADGC |
| 26449541 | 2016 | Ebbert MTW | Kauwe JSK | Interaction between variants in CLU and MS4A4E modulates Alzheimer's disease risk. | Alzheimer's & dementia : the journal of the Alzheimer's Association | 12(2):121-129 | NR | ADGC |
| 26490334 | 2015 | Escott-Price V | Williams J | Common polygenic variation enhances risk prediction for Alzheimer's disease. | Brain : a journal of neurology | 138(Pt 12):3673-84 | NR | ADGC |
| 26545630 | 2016 | Deming Y | Cruchaga C | A potential endophenotype for Alzheimer's disease: cerebrospinal fluid clusterin. | Neurobiology of aging | 37:208.e1-208.e9 | Table 3,4 | ADGC |
| 26738751 | 2017 | Ighodaro ET | Nelson PT | Risk factors and global cognitive status related to brain arteriolosclerosis in elderly individuals. | Journal of cerebral blood flow and metabolism : official journal of the International Society of Cerebral Blood Flow and Metabolism | 37(1):201-216 | NR | ADGC |
| 26827652 | 2016 | Hohman TJ | Thornton-Wells TA | Discovery of gene-gene interactions across multiple independent data sets of late onset Alzheimer disease from the Alzheimer Disease Genetics Consortium. | Neurobiology of aging | 38:141-150 | Table 2 | ADGC |
| 26830138 | 2016 | Herold C | Tanzi RE | Family-based association analyses of imputed genotypes reveal genome-wide significant association of Alzheimer's disease with OSBPL6, PTPRG, and PDCL3. | Molecular psychiatry | 21(11):1608-1612 | Table 1 | ADGC |
| 26913989 | 2016 | Traylor M | Markus HS | Shared genetic contribution to Ischaemic Stroke and Alzheimer's Disease. | Annals of neurology | 79(5):739-747 | NR | ADGC |
| 26919393 | 2016 | Karch CM | Goate AM | Alzheimer's Disease Risk Polymorphisms Regulate Gene Expression in the ZCWPW1 and the CELF1 Loci. | PloS one | 11(2):e0148717 | NR | ADGC |
| 26993346 | 2016 | Schott JM | Mead S | Genetic risk factors for the posterior cortical atrophy variant of Alzheimer's disease. | Alzheimer's & dementia : the journal of the Alzheimer's Association | 12(8):862-71 | Table 3 | ADGC |
| 27036079 | 2016 | Ridge PG | Kauwe JSK | Assessment of the genetic variance of late-onset Alzheimer's disease. | Neurobiology of aging | 41:200.e13-200.e20 | Table 1 | ADGC |
| 27066578 | 2016 | Kohli MA | Pericak-Vance MA | Segregation of a rare TTC3 variant in an extended family with late-onset Alzheimer disease. | Neurology. Genetics | 2(1):e41 | Table 1 | ADGC |
| 27088644 | 2016 | Yokoyama JS | Desikan RS | Association Between Genetic Traits for Immune-Mediated Diseases and Alzheimer Disease. | JAMA neurology | 73(6):691-7 | Table 2 | ADGC |
| 27103524 | 2016 | Mez J | Crane PK | The executive prominent/memory prominent spectrum in Alzheimer's disease is highly heritable. | Neurobiology of aging | 41:115-121 | NR | ADGC |
| 27231719 | 2016 | Cukier HN | Pericak-Vance MA | ABCA7 frameshift deletion associated with Alzheimer disease in African Americans. | Neurology. Genetics | 2(3):e79 | Table 1 | ADGC |

| Pubmed PMID | Year | First author | Last author | Title | Journal | Vol/Issue/Page | Curated tables | Curation source |
|---|---|---|---|---|---|---|---|---|
| 27357110 | 2016 | Staley LA | Kauwe JS | Genome-wide association study of prolactin levels in blood plasma and cerebrospinal fluid. | BMC genomics | 17 Suppl 3:436 | Table 1 | ADGC |
| 27357282 | 2016 | Staley LA | Kauwe JS | Variants in ACPP are associated with cerebrospinal fluid Prostatic Acid Phosphatase levels. | BMC genomics | 17 Suppl 3:439 | Table 1 | ADGC |
| 27357396 | 2016 | Ebbert MT | Kauwe JS | Variants in CCL16 are associated with blood plasma and cerebrospinal fluid CCL16 protein levels. | BMC genomics | 17 Suppl 3:437 | Table 1 | ADGC |
| 27648456 | 2016 | Bonham LW | Yokoyama JS | Age-dependent effects of APOE epsilon4 in preclinical Alzheimer's disease. | Annals of clinical and translational neurology | 3(9):668-77 | NR | ADGC |
| 27694991 | 2016 | Adams HH | Thompson PM | Novel genetic loci underlying human intracranial volume identified through genome-wide association. | Nature neuroscience | 19(12):1569-1582 | Table 1,2 | ADGC |
| 27764101 | 2016 | Jakobsdottir J | van Duijn CM | Rare Functional Variant in TM2D3 is Associated with Late-Onset Alzheimer's Disease. | PLoS genetics | 12(10):e1006327 | Table 2 | ADGC |
| 27770636 | 2017 | Mez J | Farrer LA | Two novel loci, COBL and SLC10A2, for Alzheimer's disease in African Americans. | Alzheimer's & dementia : the journal of the Alzheimer's Association | 13(2):119-129 | Table 3 | ADGC |
| 27815632 | 2016 | Nelson PT | Fardo DW | Genomics and CSF analyses implicate thyroid hormone in hippocampal sclerosis of aging. | Acta neuropathologica | 132(6):841-858 | NR | ADGC |
| 27832767 | 2016 | Deming Y | Cruchaga C | Chitinase-3-like 1 protein (CHI3L1) locus influences cerebrospinal fluid levels of YKL-40. | BMC neurology | 16(1):217 | Table 2,3 | ADGC |
| 27849641 | 2017 | Monsell SE | Kukull WA | Genetic Comparison of Symptomatic and Asymptomatic Persons With Alzheimer Disease Neuropathology. | Alzheimer disease and associated disorders | 31(3):232-238 | NR | ADGC |
| 27899424 | 2017 | Ferrari R | Desikan RS | Genetic architecture of sporadic frontotemporal dementia and overlap with Alzheimer's and Parkinson's diseases. | Journal of neurology, neurosurgery, and psychiatry | 88(2):152-164 | Table 2,3 | ADGC |
| 27933404 | 2017 | Chapuis J | Lambert JC | Genome-wide, high-content siRNA screening identifies the Alzheimer's genetic risk factor FERMT2 as a major modulator of APP metabolism. | Acta neuropathologica | 133(6):955-966 | Table 1 | ADGC |
| 27943641 | 2017 | Naj AC | Schellenberg GD | Genomic variants, genes, and pathways of Alzheimer's disease: An overview. | American journal of medical genetics. Part B, Neuropsychiatric genetics : the official publication of the International Society of Psychiatric Genetics | 174(1):5-26 | Table 2 | ADGC |
| 28106546 | 2017 | Haddick PC | van der Brug M | A Common Variant of IL-6R is Associated with Elevated IL-6 Pathway Activity in Alzheimer's Disease Brains. | Journal of Alzheimer's disease : JAD | 56(3):1037-1054 | NR | ADGC |
| 28131462 | 2017 | Katsumata Y | Fardo DW | Gene-based association study of genes linked to hippocampal sclerosis of aging neuropathology: GRN, TMEM106B, ABCC9, and KCNMB2. | Neurobiology of aging | 53:193.e17-193.e25 | Table 1,2,3,4 | ADGC |
| 28183528 | 2017 | Jun GR | Farrer LA | Transethnic genome-wide scan identifies novel Alzheimer's disease loci. | Alzheimer's & dementia : the journal of the Alzheimer's Association | 13(7):727-738 | Table 1,2 | ADGC |
| 28247064 | 2017 | Deming Y | Cruchaga C | Genome-wide association study identifies four novel loci associated with Alzheimer's endophenotypes and disease modifiers. | Acta neuropathologica | 133(5):839-856 | Table 2,3 | ADGC |
| 28323831 | 2017 | Desikan RS | Dale AM | Genetic assessment of age-associated Alzheimer disease risk: Development and validation of a polygenic hazard score. | PLoS medicine | 14(3):e1002258 | Table 2 | ADGC |

| Pubmed PMID | Year | First author | Last author | Title | Journal | Vol/Issue/Page | Curated tables | Curation source |
|---|---|---|---|---|---|---|---|---|
| 28350795 | 2017 | Steele NZ | Yokoyama JS | Fine-mapping of the human leukocyte antigen locus as a risk factor for Alzheimer disease: A case-control study. | PLoS medicine | 14(3):e1002272 | NR | ADGC |
| 28560309 | 2017 | Lee E | Zhu H | Single-nucleotide polymorphisms are associated with cognitive decline at Alzheimer's disease conversion within mild cognitive impairment patients. | Alzheimer's & dementia (Amsterdam, Netherlands) | 8:86-95 | Table 1 | ADGC |
| 28671113 | 2017 | Myrum C | Zayats T | Implication of the APP Gene in Intellectual Abilities. | Journal of Alzheimer's disease : JAD | 59(2):723-735 | NR | ADGC |
| 28714976 | 2017 | Sims R | Schellenberg GD | Rare coding variants in PLCG2, ABI3, and TREM2 implicate microglial-mediated innate immunity in Alzheimer's disease. | Nature genetics | 49(9):1373-1384 | Table 2 | ADGC |
| 28780673 | 2017 | Feng YA | Driver JA | Investigating the genetic relationship between Alzheimer's disease and cancer using GWAS summary statistics. | Human genetics | 136(10):1341-1351 | NR | ADGC |
| 28870582 | 2017 | Wang XF | Deng HW | Linking Alzheimer's disease and type 2 diabetes: Novel shared susceptibility genes detected by cFDR approach. | Journal of the neurological sciences | 380:262-272 | NR | ADGC |
| 29107063 | 2018 | Yashin AI | Ukraintseva S | Hidden heterogeneity in Alzheimer's disease: Insights from genetic association studies and other analyses. | Experimental gerontology | 107:148-160 | Table 2.1,2.2 | ADGC |
| 29274321 | 2018 | Chung J | Jun GR | Genome-wide association study of Alzheimer's disease endophenotypes at prediagnosis stages. | Alzheimer's & dementia : the journal of the Alzheimer's Association | 14(5):623-633 | Table 1,2,3 | ADGC |
| 29360470 | 2018 | Miron J | Poirier J | CDK5RAP2 gene and tau pathophysiology in late-onset sporadic Alzheimer's disease. | Alzheimer's & dementia : the journal of the Alzheimer's Association | 14(6):787-796 | Table 1 | ADGC |
| 29432188 | 2018 | Zhou X | Ip NY | Identification of genetic risk factors in the Chinese population implicates a role of immune system in Alzheimer's disease pathogenesis. | Proceedings of the National Academy of Sciences of the United States of America | 115(8):1697-1706 | NR | ADGC |
| 29458411 | 2018 | Chung J | Farrer LA | Genome-wide pleiotropy analysis of neuropathological traits related to Alzheimer's disease. | Alzheimer's research & therapy | 10(1):22 | NR | ADGC |
| 29481666 | 2018 | Teslovich TM | Mohlke KL | Identification of seven novel loci associated with amino acid levels using single-variant and gene-based tests in 8545 Finnish men from the METSIM study. | Human molecular genetics | 27(9):1664-1674 | Table 1,2 | ADGC |
| 29630712 | 2018 | Karch CM | Desikan RS | Selective Genetic Overlap Between Amyotrophic Lateral Sclerosis and Diseases of the Frontotemporal Dementia Spectrum. | JAMA neurology | 75(7):860-875 | NR | ADGC |
| 29684019 | 2018 | Lancour D | Kasif S | One for all and all for One: Improving replication of genetic studies through network diffusion. | PLoS genetics | 14(4):e1007306 | NR | ADGC |
| 29752348 | 2018 | Rutten-Jacobs LCA | Traylor M | Genetic Study of White Matter Integrity in UK Biobank (N=8448) and the Overlap With Stroke, Depression, and Dementia. | Stroke | 49(6):1340-1347 | Table 2 | ADGC |
| 29777097 | 2018 | Marioni RE | Visscher PM | GWAS on family history of Alzheimer's disease. | Translational psychiatry | 8(1):99 | NR | ADGC |
| 29801024 | 2018 | Hohman TJ | Jefferson AL | Sex-Specific Association of Apolipoprotein E With Cerebrospinal Fluid Levels of Tau. | JAMA neurology | 75(8):989-998 | Table 3 | ADGC |
| 29967939 | 2018 | Deming Y | Hohman TJ | Sex-specific genetic predictors of Alzheimer's disease biomarkers. | Acta neuropathologica | 136(6):857-872 | Table 2,3,4 | ADGC |
| 30010129 | 2018 | Ni H | Zhang C | The GWAS Risk Genes for Depression May Be Actively Involved in Alzheimer's Disease. | Journal of Alzheimer's disease : JAD | 64(4):1149-1161 | NR | ADGC |
| 30201328 | 2018 | Gusareva ES | Van Steen K | Male-specific epistasis between WWC1 and TLN2 genes is associated with Alzheimer's disease. | Neurobiology of aging | 72:188.e3-188.e12 | NR | ADGC |
| 30229991 | 2019 | Guerreiro R | Bras J | Is APOE epsilon4 required for Alzheimer's disease to develop in TREM2 p.R47H variant carriers? | Neuropathology and applied neurobiology | 45(2):187-189 | NR | ADGC |

| Pubmed PMID | Year | First author | Last author | Title | Journal | Vol/Issue/Page | Curated tables | Curation source |
|---|---|---|---|---|---|---|---|---|
| 30413934 | 2019 | Broce IJ | Desikan RS | Dissecting the genetic relationship between cardiovascular risk factors and Alzheimer's disease. | Acta neuropathologica | 137(2):209-226 | NR | ADGC |
| 30448613 | 2019 | Katsumata Y | Fardo DW | Translating Alzheimer's disease-associated polymorphisms into functional candidates: a survey of IGAP genes and SNPs. | Neurobiology of aging | 74:135-146 | Table 1,2,3 | ADGC |
| 30617256 | 2019 | Jansen IE | Posthuma D | Genome-wide meta-analysis identifies new loci and functional pathways influencing Alzheimer's disease risk. | Nature genetics | 51(3):404-413 | Table 1 | ADGC |
| 30636644 | 2019 | Nazarian A | Kulminski AM | Genome-wide analysis of genetic predisposition to Alzheimer's disease and related sex disparities. | Alzheimer's research & therapy | 11(1):5 | Table 2,3,4,5,6 | ADGC |
| 30888715 | 2019 | Lobach I | Zhang L | A simple approximation to bias in the genetic effect estimates when multiple disease states share a clinical diagnosis. | Genetic epidemiology | 43(5):522-531 | NR | ADGC |
| 30930738 | 2019 | Drange OK | Andreassen OA | Genetic Overlap Between Alzheimer's Disease and Bipolar Disorder Implicates the MARK2 and VAC14 Genes. | Frontiers in neuroscience | 0.694444444 | NR | ADGC |
| 31426376 | 2019 | Choi KY | Neuroimaging Initative AD | APOE Promoter Polymorphism-219T/G is an Effect Modifier of the Influence of APOE epsilon4 on Alzheimer's Disease Risk in a Multiracial Sample. | Journal of clinical medicine | 8(8) | Table 2,3,4 | ADGC |
| 1671712 | 1991 | Goate A | et al. | Segregation of a missense mutation in the amyloid precursor protein gene with familial Alzheimer's disease. | Nature | 349(6311):704-6 | NR | From ADGC reviews |
| 2035524 | 1991 | Pericak-Vance MA | et al. | Linkage studies in familial Alzheimer disease: evidence for chromosome 19 linkage. | American journal of human genetics | 48(6):1034-50 | NR | From ADGC reviews |
| 7596406 | 1995 | Sherrington R | St George-Hyslop PH | Cloning of a gene bearing missense mutations in early-onset familial Alzheimer's disease. | Nature | 375(6534):754-60 | NR | From ADGC reviews |
| 7638621 | 1995 | Levy-Lahad E | Schellenberg GD | A familial Alzheimer's disease locus on chromosome 1. | Science (New York, N.Y.) | 269(5226):970-3 | NR | From ADGC reviews |
| 7920638 | 1994 | Corder EH | et al. | Protective effect of apolipoprotein E type 2 allele for late onset Alzheimer disease. | Nature genetics | 7(2):180-4 | NR | From ADGC reviews |
| 9333264 | 1997 | Pericak-Vance MA | Haines JL | Complete genomic screen in late-onset familial Alzheimer disease. Evidence for a new locus on chromosome 12. | JAMA | 278(15):1237-41 | NR | From ADGC reviews |
| 11113612 | 2000 | Pericak-Vance MA | Haines JL | Identification of novel genes in late-onset Alzheimer's disease. | Experimental gerontology | 35(9-10):1343-52 | NR | From ADGC reviews |
| 11806855 | 2001 | Curtis D | Sham PC | A novel method of two-locus linkage analysis applied to a genome scan for late onset Alzheimer's disease. | Annals of human genetics | 65(Pt 5):473-81 | NR | From ADGC reviews |
| 11857588 | 2002 | Myers A | Goate A | Full genome screen for Alzheimer disease: stage II analysis. | American journal of medical genetics | 114(2):235-44 | NR | From ADGC reviews |
| 11875758 | 2002 | Li YJ | Pericak-Vance MA | Age at onset in two common neurodegenerative diseases is genetically controlled. | American journal of human genetics | 70(4):985-93 | NR | From ADGC reviews |
| 12016588 | 2002 | Olson JM | Dudek DM | A second locus for very-late-onset Alzheimer disease: a genome scan reveals linkage to 20p and epistasis between 20p and the amyloid precursor protein region. | American journal of human genetics | 71(1):154-61 | NR | From ADGC reviews |
| 12490529 | 2003 | Blacker D | Tanzi RE | Results of a high-resolution genome screen of 437 Alzheimer's disease families. | Human molecular genetics | 12(1):23-32 | NR | From ADGC reviews |
| 14564669 | 2003 | Scott WK | Pericak-Vance MA | Ordered-subsets linkage analysis detects novel Alzheimer disease loci on chromosomes 2q34 and 15q22. | American journal of human genetics | 73(5):1041-51 | NR | From ADGC reviews |

| Pubmed PMID | Year | First author | Last author | Title | Journal | Vol/Issue/Page | Curated tables | Curation source |
|---|---|---|---|---|---|---|---|---|
| 15729734 | 2005 | Holmans P | Williams J | Genome screen for loci influencing age at onset and rate of decline in late onset Alzheimer's disease. | American journal of medical genetics. Part B, Neuropsychiatric genetics : the official publication of the International Society of Psychiatric Genetics | 135B(1):24-32 | NR | From ADGC reviews |
| 17101828 | 2006 | Lee JH | Mayeux R | Expanded genomewide scan implicates a novel locus at 3q28 among Caribbean hispanics with familial Alzheimer disease. | Archives of neurology | 63(11):1591-8 | NR | From ADGC reviews |
| 17317784 | 2007 | Grupe A | Williams J | Evidence for novel susceptibility genes for late-onset Alzheimer's disease from a genome-wide association study of putative functional variants. | Human molecular genetics | 16(8):865-73 | Table 2,4 | From ADGC reviews |
| 17564960 | 2007 | Liu F | van Duijn CM | A genomewide screen for late-onset Alzheimer disease in a genetically isolated Dutch population. | American journal of human genetics | 81(1):17-31 | Table 6 | From ADGC reviews |
| 17725986 | 2007 | Hamshere ML | Owen MJ | Genome-wide linkage analysis of 723 affected relative pairs with late-onset Alzheimer's disease. | Human molecular genetics | 16(22):2703-12 | NR | From ADGC reviews |
| 17940814 | 2008 | Lee JH | Mayeux R | Age-at-onset linkage analysis in Caribbean Hispanics with familial late-onset Alzheimer's disease. | Neurogenetics | 9(1):51-60 | NR | From ADGC reviews |
| 17957224 | 2008 | Sillen A | Graff C | Expanded high-resolution genetic study of 109 Swedish families with Alzheimer's disease. | European journal of human genetics : EJHG | 16(2):202-8 | NR | From ADGC reviews |
| 17474819 | 2007 | Coon KD | Stephan DA | A high-density whole-genome association study reveals that APOE is the major susceptibility gene for sporadic late-onset Alzheimer's disease. | The Journal of clinical psychiatry | 68(4):613-8 | NR | GWAS catalog |
| 17553421 | 2007 | Reiman EM | Stephan DA | GAB2 alleles modify Alzheimer's risk in APOE epsilon4 carriers. | Neuron | 54(5):713-20 | Table 1 | GWAS catalog |
| 17975299 | 2008 | Webster JA | Stephan DA | Sorl1 as an Alzheimer's disease predisposition gene? | Neuro-degenerative diseases | 5(2):60-4 | Table 1 | GWAS catalog |
| 17998437 | 2008 | Li H | Roses AD | Candidate single-nucleotide polymorphisms from a genomewide association study of Alzheimer disease. | Archives of neurology | 65(1):45-53 | Table 2,3 | GWAS catalog |
| 18449908 | 2009 | Poduslo SE | Smith S | Genome screen of late-onset Alzheimer's extended pedigrees identifies TRPC4AP by haplotype analysis. | American journal of medical genetics. Part B, Neuropsychiatric genetics : the official publication of the International Society of Psychiatric Genetics | 150B(1):50-5 | Table 1 | GWAS catalog |
| 18823527 | 2008 | Abraham R | Kirov G | A genome-wide association study for late-onset Alzheimer's disease using DNA pooling. | BMC medical genomics | 1:44 | Table 1,2 | GWAS catalog |
| 18976728 | 2008 | Bertram L | Tanzi RE | Genome-wide association analysis reveals putative Alzheimer's disease susceptibility loci in addition to APOE. | American journal of human genetics | 83(5):623-32 | Table 1,2 | GWAS catalog |
| 19118814 | 2009 | Beecham GW | Pericak-Vance MA | Genome-wide association study implicates a chromosome 12 risk locus for late-onset Alzheimer disease. | American journal of human genetics | 84(1):35-43 | Table 2,3,5 | GWAS catalog |
| 19125160 | 2010 | Feulner TM | Riemenschneider M | Examination of the current top candidate genes for AD in a genome-wide association study. | Molecular psychiatry | 15(7):756-66 | NR | GWAS catalog |
| 19136949 | 2009 | Carrasquillo MM | Younkin SG | Genetic variation in PCDH11X is associated with susceptibility to late-onset Alzheimer's disease. | Nature genetics | 41(2):192-8 | Table 1,2,4,5 | GWAS catalog |
| 19362756 | 2009 | Butler AW | Powell JF | Meta-analysis of linkage studies for Alzheimer's disease--a web resource. | Neurobiology of aging | 30(7):1037-47 | NR | GWAS catalog |
| 19734902 | 2009 | Harold D | Williams J | Genome-wide association study identifies variants at CLU and PICALM associated with Alzheimer's disease. | Nature genetics | 41(10):1088-93 | Table 1,2 | GWAS catalog |

| Pubmed PMID | Year | First author | Last author | Title | Journal | Vol/Issue/Page | Curated tables | Curation source |
|---|---|---|---|---|---|---|---|---|
| 19734903 | 2009 | Lambert JC | Amouyel P | Genome-wide association study identifies variants at CLU and CR1 associated with Alzheimer's disease. | Nature genetics | 41(10):1094-9 | Table 1,3 | GWAS catalog |
| 20061627 | 2010 | Heinzen EL | Goldstein DB | Genome-wide scan of copy number variation in late-onset Alzheimer's disease. | Journal of Alzheimer's disease : JAD | 19(1):69-77 | Table 2 | GWAS catalog |
| 20197096 | 2010 | Stein JL | Thompson PM | Genome-wide analysis reveals novel genes influencing temporal lobe structure with relevance to neurodegeneration in Alzheimer's disease. | NeuroImage | 51(2):542-54 | Table 1 | GWAS catalog |
| 20452100 | 2011 | Kramer PL | Ott J | Alzheimer disease pathology in cognitively healthy elderly: a genome-wide study. | Neurobiology of aging | 32(12):2113-22 | Table 2 | GWAS catalog |
| 20460622 | 2010 | Seshadri S | Breteler MM | Genome-wide analysis of genetic loci associated with Alzheimer disease. | JAMA | 303(18):1832-40 | Table 2,3 | GWAS catalog |
| 20534741 | 2010 | Corneveaux JJ | Huentelman MJ | Association of CR1, CLU and PICALM with Alzheimer's disease in a cohort of clinically characterized and neuropathologically verified individuals. | Human molecular genetics | 19(16):3295-301 | Table 2 | GWAS catalog |
| 20558387 | 2010 | Biffi A | Rosand J | Genetic variation and neuroimaging measures in Alzheimer disease. | Archives of neurology | 67(6):677-85 | Table 4,5 | GWAS catalog |
| 20885792 | 2010 | Naj AC | Pericak-Vance MA | Dementia revealed: novel chromosome 6 locus for late-onset Alzheimer disease provides genetic evidence for folate-pathway abnormalities. | PLoS genetics | 6(9):e1001130 | Table 2 | GWAS catalog |
| 21059989 | 2011 | Lee JH | Mayeux R | Identification of novel loci for Alzheimer disease and replication of CLU, PICALM, and BIN1 in Caribbean Hispanic individuals. | Archives of neurology | 68(3):320-8 | NR | GWAS catalog |
| 21098978 | 2011 | Sherva R | Farrer LA | Identification of novel candidate genes for Alzheimer's disease by autozygosity mapping using genome wide SNP data. | Journal of Alzheimer's disease : JAD | 23(2):349-59 | Table 3,4 | GWAS catalog |
| 21116278 | 2011 | Furney SJ | Lovestone S | Genome-wide association with MRI atrophy measures as a quantitative trait locus for Alzheimer's disease. | Molecular psychiatry | 16(11):1130-8 | Table 2 | GWAS catalog |
| 21220680 | 2011 | Reitz C | Mayeux R | Meta-analysis of the association between variants in SORL1 and Alzheimer disease. | Archives of neurology | 68(1):99-106 | Table 3,4,5 | GWAS catalog |
| 21379329 | 2011 | Wijsman EM | Mayeux R | Genome-wide association of familial late-onset Alzheimer's disease replicates BIN1 and CLU and nominates CUGBP2 in interaction with APOE. | PLoS genetics | 7(2):e1001308 | Table 4,5 | GWAS catalog |
| 21390209 | 2011 | Hu X | Soares H | Meta-analysis for genome-wide association study identifies multiple variants at the BIN1 locus associated with late-onset Alzheimer's disease. | PloS one | 6(2):e16616 | Table 1,2,3,5 | GWAS catalog |
| 21627779 | 2011 | Antunez C | Ruiz A | The membrane-spanning 4-domains, subfamily A (MS4A) gene cluster contains a common variant associated with Alzheimer's disease. | Genome medicine | 3(5):33 | NR | GWAS catalog |
| 22005930 | 2012 | Hollingworth P | Williams J | Genome-wide association study of Alzheimer's disease with psychotic symptoms. | Molecular psychiatry | 17(12):1316-27 | Table 2,3 | GWAS catalog |
| 22005931 | 2012 | Kamboh MI | Lopez OL | Genome-wide association analysis of age-at-onset in Alzheimer's disease. | Molecular psychiatry | 17(12):1340-6 | Table 1 | GWAS catalog |
| 22159054 | 2011 | Logue MW | Farrer LA | A comprehensive genetic association study of Alzheimer disease in African Americans. | Archives of neurology | 68(12):1569-79 | Table 4,5,6 | GWAS catalog |
| 22430674 | 2013 | Lambert JC | Amouyel P | Genome-wide haplotype association study identifies the FRMD4A gene as a risk locus for Alzheimer's disease. | Molecular psychiatry | 18(4):461-70 | NR | GWAS catalog |

| Pubmed PMID | Year | First author | Last author | Title | Journal | Vol/Issue/Page | Curated tables | Curation source |
|---|---|---|---|---|---|---|---|---|
| 22539578 | 2012 | Barral S | Mayeux R | Genotype patterns at PICALM, CR1, BIN1, CLU, and APOE genes are associated with episodic memory. | Neurology | 78(19):1464-71 | Table 3,4 | GWAS catalog |
| 22556362 | 2012 | Coppola G | Geschwind DH | Evidence for a role of the rare p.A152T variant in MAPT in increasing the risk for FTD-spectrum and Alzheimer's diseases. | Human molecular genetics | 21(15):3500-12 | NR | GWAS catalog |
| 22785395 | 2012 | Gaj P | Ostrowski J | Identification of a late onset Alzheimer's disease candidate risk variant at 9q21.33 in Polish patients. | Journal of Alzheimer's disease : JAD | 32(1):157-68 | NR | GWAS catalog |
| 22801501 | 2012 | Jonsson T | Stefansson K | A mutation in APP protects against Alzheimer's disease and age-related cognitive decline. | Nature | 488(7409):96-9 | NR | GWAS catalog |
| 22832961 | 2012 | Kamboh MI | Barmada MM | Genome-wide association study of Alzheimer's disease. | Translational psychiatry | 2:e117 | Table 1 | GWAS catalog |
| 22881374 | 2012 | Cummings AC | Haines JL | Genome-wide association and linkage study in the Amish detects a novel candidate late-onset Alzheimer disease gene. | Annals of human genetics | 76(5):342-51 | Table 4 | GWAS catalog |
| 23150908 | 2013 | Jonsson T | Stefansson K | Variant of TREM2 associated with the risk of Alzheimer's disease. | The New England journal of medicine | 368(2):107-16 | Table 1,2 | GWAS catalog |
| 23150934 | 2013 | Guerreiro R | Hardy J | TREM2 variants in Alzheimer's disease. | The New England journal of medicine | 368(2):117-27 | Table 2 | GWAS catalog |
| 23374588 | 2013 | Martinelli-Boneschi F | Albani D | Pharmacogenomics in Alzheimer's disease: a genome-wide association study of response to cholinesterase inhibitors. | Neurobiology of aging | 34(6):1711.e7-13 | Table 2 | GWAS catalog |
| 23419831 | 2014 | Ramanan VK | Saykin AJ | APOE and BCHE as modulators of cerebral amyloid deposition: a florbetapir PET genome-wide association study. | Molecular psychiatry | 19(3):351-7 | NR | GWAS catalog |
| 23541187 | 2014 | Swaminathan S | Saykin AJ | Association of plasma and cortical amyloid beta is modulated by APOE epsilon4 status. | Alzheimer's & dementia : the journal of the Alzheimer's Association | 10(1):e9-e18 | Table 1,2 | GWAS catalog |
| 24958192 | 2014 | Gusareva ES | Van Steen K | Genome-wide association interaction analysis for Alzheimer's disease. | Neurobiology of aging | 35(11):2436-2443 | Table 2 | GWAS catalog |
| 25027320 | 2014 | Ramirez A | Nothen MM | SUCLG2 identified as both a determinator of CSF Abeta1-42 levels and an attenuator of cognitive decline in Alzheimer's disease. | Human molecular genetics | 23(24):6644-58 | Table 2,3,4 | GWAS catalog |
| 25311924 | 2014 | Chouraki V | Seshadri S | Genetics of Alzheimer's disease. | Advances in genetics | 87:245-94 | NR | GWAS catalog |
| 30651383 | 2019 | Chauhan G | Debette S | Genetic and lifestyle risk factors for MRI-defined brain infarcts in a population-based setting. | Neurology | | Table 1,2,3,4 | GWAS catalog |
| 30805717 | 2019 | Zhu Z | Liang L | Shared genetic architecture between metabolic traits and Alzheimer's disease: a large-scale genome-wide cross-trait analysis. | Human genetics | 138(3):271-285 | NR | GWAS catalog |
| 30820047 | 2019 | Kunkle BW | Pericak-Vance MA | Genetic meta-analysis of diagnosed Alzheimer's disease identifies new risk loci and implicates Abeta, tau, immunity and lipid processing. | Nature genetics | 51(3):414-430 | Table 1,2 | GWAS catalog |
| 30979435 | 2019 | Lo MT | Chen CH | Identification of genetic heterogeneity of Alzheimer's disease across age. | Neurobiology of aging | 84:243.e1-243.e9 | NR | GWAS catalog |
| 31055733 | 2019 | Nazarian A | Kulminski AM | Genetic heterogeneity of Alzheimer's disease in subjects with and without hypertension. | GeroScience | 41(2):137-154 | Table 1,2,3,4,5 | GWAS catalog |
| 31497858 | 2019 | Dumitrescu L | Hohman TJ | Sex differences in the genetic predictors of Alzheimer's pathology. | Brain : a journal of neurology | 142(9):2581-2589 | Table 1,2 | GWAS catalog |

| Pubmed PMID | Year | First author | Last author | Title | Journal | Vol/Issue/Page | Curated tables | Curation source |
|---|---|---|---|---|---|---|---|---|
| 20932310 | 2010 | Han MR | Wang LS | Genome-wide association reveals genetic effects on human Abeta42 and tau protein levels in cerebrospinal fluids: a case control study. | BMC neurology | 0.479166667 | NR | GWAS catalog, ADGC |
| 21123754 | 2011 | Kim S | Saykin AJ | Genome-wide association study of CSF biomarkers Abeta1-42, t-tau, and p-tau181p in the ADNI cohort. | Neurology | 76(1):69-79 | Table 2 | GWAS catalog, ADGC |
| 21312009 | 2011 | Sherva R | Farrer LA | Power and pitfalls of the genome-wide association study approach to identify genes for Alzheimer's disease. | Current psychiatry reports | 13(2):138-46 | NR | GWAS catalog, ADGC |
| 21460840 | 2011 | Hollingworth P | Williams J | Common variants at ABCA7, MS4A6A/MS4A4E, EPHA1, CD33 and CD2AP are associated with Alzheimer's disease. | Nature genetics | 43(5):429-35 | Table 1,2 | GWAS catalog, ADGC |
| 21901424 | 2012 | Swaminathan S | Saykin AJ | Amyloid pathway-based candidate gene analysis of [(11)C]PiB-PET in the Alzheimer's Disease Neuroimaging Initiative (ADNI) cohort. | Brain imaging and behavior | 6(1):1-15 | Table 1,3,4 | GWAS catalog, ADGC |
| 22245343 | 2012 | Meda SA | Pearlson GD | A large scale multivariate parallel ICA method reveals novel imaging-genetic relationships for Alzheimer's disease in the ADNI cohort. | NeuroImage | 60(3):1608-21 | Table 3 | GWAS catalog, ADGC |
| 22343898 | 2012 | Liu N | Bonini NM | The microRNA miR-34 modulates ageing and neurodegeneration in Drosophila. | Nature | 482(7386):519-23 | NR | GWAS catalog, ADGC |
| 22431837 | 2012 | Galasko DR | Aisen P | Antioxidants for Alzheimer disease: a randomized clinical trial with cerebrospinal fluid biomarker measures. | Archives of neurology | 69(7):836-41 | NR | GWAS catalog, ADGC |
| 22480918 | 2012 | Ramanan VK | Saykin AJ | Pathway analysis of genomic data: concepts, methods, and prospects for future development. | Trends in genetics : TIG | 28(7):323-32 | NR | GWAS catalog, ADGC |
| 22618995 | 2012 | Schellenberg GD | Montine TJ | The genetics and neuropathology of Alzheimer's disease. | Acta neuropathologica | 124(3):305-23 | Table 2 | GWAS catalog, ADGC |
| 22673115 | 2012 | Vardarajan BN | Farrer LA | Identification of Alzheimer disease-associated variants in genes that regulate retromer function. | Neurobiology of aging | 33(9):2231.e15-2231.e30 | Table 3 | GWAS catalog, ADGC |
| 22685416 | 2012 | Zou F | Ertekin-Taner N | Brain expression genome-wide association study (eGWAS) identifies human disease-associated variants. | PLoS genetics | 8(6):e1002707 | Table 1,3,5 | GWAS catalog, ADGC |
| 22722634 | 2012 | Allen M | Woltjer RL | Novel late-onset Alzheimer disease loci variants associate with brain gene expression. | Neurology | 79(3):221-8 | Table 2,3 | GWAS catalog, ADGC |
| 22821396 | 2012 | Cruchaga C | Goate AM | Cerebrospinal fluid APOE levels: an endophenotype for genetic studies for Alzheimer's disease. | Human molecular genetics | 21(20):4558-71 | NR | GWAS catalog, ADGC |
| 22869155 | 2012 | Jun G | Farrer LA | Comprehensive search for Alzheimer disease susceptibility loci in the APOE region. | Archives of neurology | 69(10):1270-9 | Table 2,3,4 | GWAS catalog, ADGC |
| 22903471 | 2013 | Hibar DP | Thompson PM | Genome-wide association identifies genetic variants associated with lentiform nucleus volume in N = 1345 young and elderly subjects. | Brain imaging and behavior | 7(2):102-15 | Table 2 | GWAS catalog, ADGC |
| 23107433 | 2013 | Kohli MA | Zuchner S | Repeat expansions in the C9ORF72 gene contribute to Alzheimer's disease in Caucasians. | Neurobiology of aging | 34(5):1519.e5-12 | NR | GWAS catalog, ADGC |
| 23143602 | 2012 | Whitcomb DC | Devlin B | Common genetic variants in the CLDN2 and PRSS1-PRSS2 loci alter risk for alcohol-related and sporadic pancreatitis. | Nature genetics | 44(12):1349-54 | Table 2 | GWAS catalog, ADGC |
| 23360175 | 2013 | Holton P | Guerreiro R | Initial assessment of the pathogenic mechanisms of the recently identified Alzheimer risk Loci. | Annals of human genetics | 77(2):85-105 | Table 1 | GWAS catalog, ADGC |
| 23535033 | 2014 | Sherva R | Green RC | Genome-wide association study of the rate of cognitive decline in Alzheimer's disease. | Alzheimer's & dementia : the journal of the Alzheimer's Association | 10(1):45-52 | Table 2 | GWAS catalog, ADGC |

| Pubmed PMID | Year | First author | Last author | Title | Journal | Vol/Issue/Page | Curated tables | Curation source |
|---|---|---|---|---|---|---|---|---|
| 23562540 | 2013 | Cruchaga C | Goate AM | GWAS of cerebrospinal fluid tau levels identifies risk variants for Alzheimer's disease. | Neuron | 78(2):256-68 | Table 2,5 | GWAS catalog, ADGC |
| 24922517 | 2014 | Escott-Price V | Williams J | Gene-wide analysis detects two new susceptibility genes for Alzheimer's disease. | PloS one | 9(6):e94661 | Table 3 | GWAS catalog, ADGC |
| 25043464 | 2014 | Jun G | Farrer LA | PLXNA4 is associated with Alzheimer disease and modulates tau phosphorylation. | Annals of neurology | 76(3):379-92 | Table 2 | GWAS catalog, ADGC |
| 25150575 | 2014 | Barral S | Mayeux R | Genetic variants in a 'cAMP element binding protein' (CREB)-dependent histone acetylation pathway influence memory performance in cognitively healthy elderly individuals. | Neurobiology of aging | 35(12):2881.e7-2881.e10 | Table 1,2 | GWAS catalog, ADGC |
| 25172201 | 2014 | Logue MW | Manly JJ | Two rare AKAP9 variants are associated with Alzheimer's disease in African Americans. | Alzheimer's & dementia : the journal of the Alzheimer's Association | 10(6):609-618.e11 | Table 1,2 | GWAS catalog, ADGC |
| 25188341 | 2014 | Beecham GW | Montine TJ | Genome-wide association meta-analysis of neuropathologic features of Alzheimer's disease and related dementias. | PLoS genetics | 10(9):e1004606 | Table 2,3 | GWAS catalog, ADGC |
| 25199842 | 2014 | Naj AC | Yu L | Effects of multiple genetic loci on age at onset in late-onset Alzheimer disease: a genome-wide association study. | JAMA neurology | 71(11):1394-404 | Table 1 | GWAS catalog, ADGC |
| 25317765 | 2014 | Barral S | Mayeux R | Common genetic variants on 6q24 associated with exceptional episodic memory performance in the elderly. | JAMA neurology | 71(12):1514-9 | NR | GWAS catalog, ADGC |
| 25324900 | 2014 | Allen M | Ertekin-Taner N | Association of MAPT haplotypes with Alzheimer's disease risk and MAPT brain gene expression levels. | Alzheimer's research & therapy | 6(4):39 | Table 1,3 | GWAS catalog, ADGC |
| 25340798 | 2014 | Kauwe JS | Goate AM | Genome-wide association study of CSF levels of 59 alzheimer's disease candidate proteins: significant associations with proteins involved in amyloid processing and inflammation. | PLoS genetics | 10(10):e1004758 | Table 2 | GWAS catalog, ADGC |

### Supplementary Table 2 – All curated and harmonized/derived data fields in ADVP

| Column index | Column names | How the information was obtained | Description of the column |
|---|---|---|---|
| 1 | Pubmed ID | Extract | Pubmed PMID of the publication |
| 2 | Record Type | Infer | SNP-based or Gene-based association |
| 3 | SNP | Extract | rsID shown in the publication |
| 4 | Coordinates | Extract | Chromosome and base-pair informaton shown in publication (note can vary across genome builds) |
| 5 | Locus | Extract | Reported gene in the association record in the results table in the publication (not the eQTL or gene-based gene) |
| 6 | Reported gene | Extract | Gene name for gene-based test. NA if that's SNP-based test. |
| 7 | Interactions | Extract | Reported interaction results |
| 8 | Population | Infer | Choose from the following, derived from "Population" column, population mapped into these categories: African American, Arab, Asian, Carribbean Hispanic, Caucasian, Hispanic, Non-Hispanic Caucasian, Non-Hispanic White |
| 9 | Population (detailed) | Extract | Population description from the publication |
| 10 | Cohort | Infer | Cohorts derived from Cohort (detailed) column: if consortium name was available (e.g. ADGC, CHARGE, these will be used) |
| 11 | Cohort (detailed) | Extract | Cohort description from the publication |
| 12 | Sample size | Extract | Total sample size |
| 13 | Subset Analyzed | Extract | What samples were used for analyses. This is designed for distinguishing between same snps with different p-values in the same data table in a publication. |
| 14 | Phenotype | Infer | Derived from Phenotype (detailed) column. Choose from the following: AD, ADRD, Cognitive, Expression, Fluid biomarker, Imaging, Neuropathology, Non-ADRD, Other |
| 15 | Phenotype (detailed) | Extract | Defined as outcome of the regression analyses, shown is what is described in text |
| 16 | Association type | Infer | Choose from the following: eQTL, Disease risk, Endophenotype, AAO/Survival, Pleiotropy, Cross phenotype |
| 17 | RA1 | Extract | Reported Allele 1 - the first allele reported in the association record in the publication, if any |
| 18 | RA2 | Extract | Reported Allele 2 - the second allele reported in the association record in the publication, if any |
| 19 | AF | Extract | Reported allele frequency across all samples (for this associationr record) in the publication |
| 20 | P-value | Extract | p-value reported; show corrected p-value if available |
| 21 | Effect Size | Extract | Effect size type (OR, Beta, FDR etc) and value of the effect size |
| 22 | Confidence Interval | Extract | 95% confidence interval of the effect size (if any) |
| 23 | Stage | Infer | Stage of the analysis reported in the publication, e.g., “Stage n” (n=1,2,3). If nothing is reported, choose from the followings: “Discovery”, “Validation”, “Meta-analysis", "Joint-analysis" |
| 24 | Model | Extract | Description of model: what kind of statistical model, and if the analyses were adjusted for anything |

| Column index | Column names | How the information was obtained | Description of the column |
|---|---|---|---|
| 25 | Imputation | Extract | How the data is imputed. Choose from the following: 1000G, HapMap, HRC |
| 26 | View in GenomicsDB | Compute / cross-reference | URL link for viewing the record in NIAGADS genomicsDB |
| 27 | Nearest gene | Compute | Distance from the SNP to the Locus (basepair information included) |
| 28 | Most severe consequence | Compute / cross-reference | Functional information (VEP provided by NIAGADS genomicDB) |

### Supplementary Table 3 – ADGC cohorts included in ADVP

| Dataset | Full name | Race/Ethnicity |
|---|---|---|
| ACT1 (includes GeneticDifferences) | Adult Changes in Thought | Caucasian |
| ADC1 | NIA Alzheimer's Disease Centers | Caucasian |
| ADC2 | NIA Alzheimer's Disease Centers | Caucasian |
| ADC3 | NIA Alzheimer's Disease Centers | Caucasian |
| ADC4 | NIA Alzheimer's Disease Centers | Caucasian |
| ADC5 | NIA Alzheimer's Disease Centers | Caucasian |
| ADC6 | NIA Alzheimer's Disease Centers | Caucasian |
| ADC7 | NIA Alzheimer's Disease Centers | Caucasian |
| ADNI | Alzheimer's Disease Neuroimaging Initiative | Caucasian |
| BIOCARD | Biomarkers of Cognitive Decline Among Normal Individuals | Caucasian |
| CHAP | Chicago Health and Aging Project | Caucasian |
| EAS | Einstein Aging Study | Caucasian |
| GSK | Genotype-Phenotype Associations in Alzheimer's Disease | Caucasian |
| NIA-LOAD | NIA-Late Onset of Alzheimer's Disease/National Centralized Repository for Alzheimer's Disease | Caucasian |
| MAYO | Mayo Clinic | Caucasian |
| MIRAGE | Multi-Institutional Research in Alzheimer's Genetic Epidemiology | Caucasian |
| NBB | Netherlands Brain Bank | Caucasian |
| OHSU | Oregon Health and Science University | Caucasian |
| PFIZER | Pfizer Lipitor’s Effect in Alzheimer’s Disease (LEADe) trial/PrecisionMed case-control study/Phas | Caucasian |
| RMAYO | Rochester Mayo Clinic | Caucasian |
| ROSMAP1 | Religious Orders Study / Memory and Aging Project | Caucasian |
| ROSMAP2 | Religious Orders Study / Memory and Aging Project | Caucasian |
| TARC1 | Texas Alzheimer's Research and Care Consortium | Caucasian |
| TGEN2 | Translational Genomics Research Institute Series 2 | Caucasian |
| UKS | Universitat Saarlandes | Caucasian |
| UM/VU/MSSM | University of Miami/Vanderbilt University University/Mt. Sinai School of Medicine | Caucasian |
| UM/VU/MSSM | University of Miami/Vanderbilt University University/Mt. Sinai School of Medicine | Caucasian |
| UM/VU/MSSM | University of Miami/Vanderbilt University University/Mt. Sinai School of Medicine | Caucasian |
| UM/VU/TARC2 (MTV/MTC) | University of Miami/Vanderbilt University University/Texas Alzheimer's Research and Care Consortium | Caucasian |
| UPITT | University of Pittsburgh | Caucasian |
| WASHU1 | Washington University | Caucasian |
| WASHU2 | Washington University | Caucasian |

| Dataset | Full name | Race/Ethnicity |
|---|---|---|
| WHICAP | Washington Heights/Inwood Columbia Aging Project | Caucasian |
| ACT | Adult Changes in Thought | African American |
| ADC1/2 | NIA Alzheimer's Disease Centers | African American |
| ADC3 | NIA Alzheimer's Disease Centers | African American |
| ADC8_AA | NIA Alzheimer's Disease Centers | African American |
| CHOP_AA (ADGC (2013*)) | See below* | African American |
| CHOP_AA Redos (ADGC (2018a)‡) | See below‡ | African American |
| ADC9_AA (ADGC 2018b) | NIA Alzheimer's Disease Centers | African American |
| CHAP | Chicago Health and Aging Project | African American |
| GenerAAtions | Genetic and Environmental Risk Factors for Alzheimer’s Disease Among African Americans | African American |
| Indianapolis | Indianapolis African Americans/Ibadan Study of Aging | African American |
| Mirage 300 | Multi-Institutional Research in Alzheimer's Genetic Epidemiology | African American |
| Mirage 660 | Multi-Institutional Research in Alzheimer's Genetic Epidemiology | African American |
| NIA-LOAD/NCRAD | NIA-Late Onset of Alzheimer's Disease/National Centralized Repository for Alzheimer's Disease | African American |
| GARD Study | Gwangju Alzheimer and Related Dementias Study (Kunho Lee) | Asian |
| JGSCAD | Japanese Genetic Study Consortium of Alzheimer's Disease (Ryozo Kuwano, Niigata Univ) | Asian |
| Korean | Samsung Medical Center Korea (Jong-Won Kim) | Asian |

*Cohorts included in CHOP_AA (ADGC (2013))

‡ Cohorts included in CHOP_AA Redos (ADGC (2018a))

**Supplementary Table 4 – Top AD-associated loci and SNPs across populations**

| Reported locus (nearest gene) | AfricanAmerican_TopSNP | AfricanAmerican_OR | AfricanAmerican_P-value | AfricanAmerican_MAF | Asian_TopSNP | Asian_OR | Asian_P-value | Asian_MAF | CaribbeanHispanic_TopSNP | CaribbeanHispanic_OR | CaribbeanHispanic_P-value | CaribbeanHispanic_MAF | NHW_TopSNP | NHW_OR | NHW_P-value | NHW_MAF |
|---|---|---|---|---|---|---|---|---|---|---|---|---|---|---|---|---|
| ABCA7 | rs115550680 | 1.79 | 2.21E-09 | 0.07 | NA | NA | NA | NA | NA | NA | NA | NA | rs3752246 | 1.15 | 3.10E-16 | 0.182 |
| AC098650.1 | rs2221154 | 0.57 | 2.58E-06 | 0.19 | NA | NA | NA | NA | NA | NA | NA | NA | rs6504163 | 0.952381 | 5.40E-09 | 0.37 |
| ACE | NA | NA | NA | NA | NA | NA | NA | NA | NA | NA | NA | NA | rs593742 | 0.943396 | 6.20E-11 | 0.31 |
| ADAM10 | NA | NA | NA | NA | NA | NA | NA | NA | NA | NA | NA | NA | rs2830500 | 0.93 | 2.60E-08 | 0.308 |
| ADAMTS1 | NA | NA | NA | NA | NA | NA | NA | NA | NA | NA | NA | NA | rs7295246 | 1.07 | 2.70E-07 | 0.413 |
| ADAMTS20 | NA | NA | NA | NA | NA | NA | NA | NA | NA | NA | NA | NA | rs4296166 | 1.12 | 4.08E-06 | 0.478 |
| AKAP6 | NA | NA | NA | NA | NA | NA | NA | NA | NA | NA | NA | NA | rs445925 | 0.74 | 4.10E-11 | 0.12 |
| ARHGAP45 | rs115553053 | 1.86 | 3.14E-08 | 0.06 | NA | NA | NA | NA | NA | NA | NA | NA | NA | NA | NA | NA |
| ARID5B | NA | NA | NA | NA | NA | NA | NA | NA | NA | NA | NA | NA | rs2588969 | 0.88 | 6.90E-07 | 0.366 |
| ARL17B | NA | NA | NA | NA | NA | NA | NA | NA | NA | NA | NA | NA | rs2732703 | 0.73 | 5.80E-09 | 0.13 |
| BCAM | NA | NA | NA | NA | NA | NA | NA | NA | NA | NA | NA | NA | rs10402271 | 1.26 | 2.14E-07 | 0.36 |
| BCKDK | NA | NA | NA | NA | NA | NA | NA | NA | NA | NA | NA | NA | rs889555 | 0.95 | 3.20E-08 | 0.29 |
| BCL3 | NA | NA | NA | NA | NA | NA | NA | NA | NA | NA | NA | NA | rs2965101 | 0.934579 | 5.50E-03 | 0.31 |
| BIN1 | rs11685593 | 1.66 | 9.80E-03 | 0.06 | rs744373 | 1.25 | 1.39E-04 | 0.33 | NA | NA | NA | NA | rs9331896 | 0.88 | 4.60E-24 | 0.387 |
| CACNA2D3 | NA | NA | NA | NA | NA | NA | NA | NA | rs7431992 | 1.59 | 1.99E-08 | 0.1 | NA | NA | NA | NA |
| CASS4 | NA | NA | NA | NA | NA | NA | NA | NA | NA | NA | NA | NA | rs6024870 | 0.88 | 3.50E-08 | 0.088 |
| CCDC83 | NA | NA | NA | NA | rs10898417 | 0.59 | 1.17E-06 | 0.15 | NA | NA | NA | NA | NA | NA | NA | NA |
| CD2AP | NA | NA | NA | NA | NA | NA | NA | NA | NA | NA | NA | NA | rs9473117 | 1.09 | 1.20E-10 | 0.28 |
| CD33 | NA | NA | NA | NA | NA | NA | NA | NA | rs3865444 | 0.87 | 8.00E-03 | 0.25 | rs3865444 | 0.91 | 5.10E-08 | 0.307 |
| CDC42SE2 | NA | NA | NA | NA | NA | NA | NA | NA | NA | NA | NA | NA | rs382216 | 0.87 | 2.00E-07 | 0.36 |
| CDH9 | NA | NA | NA | NA | NA | NA | NA | NA | NA | NA | NA | NA | rs71618613 | 0.71 | 3.30E-07 | 0.01 |
| CEACAM16 | NA | NA | NA | NA | NA | NA | NA | NA | NA | NA | NA | NA | rs2965109 | 0.92 | 2.70E-03 | 0.38 |
| CELF1 | NA | NA | NA | NA | NA | NA | NA | NA | NA | NA | NA | NA | rs10838725 | 1.08 | 6.70E-06 | 0.316 |
| CLDN18 | NA | NA | NA | NA | NA | NA | NA | NA | NA | NA | NA | NA | rs16847609 | 1.19 | 5.30E-07 | 0.09 |
| CLU | NA | NA | NA | NA | rs2279590 | 0.85 | 7.01E-03 | 0.25 | NA | NA | NA | NA | rs11218343 | 0.8 | 2.90E-12 | 0.04 |
| CNTNAP2 | rs10273775 | 1.52 | 8.94E-06 | 0.42 | NA | NA | NA | NA | NA | NA | NA | NA | NA | NA | NA | NA |
| COBL | rs112404845 | 3.59 | 8.70E-07 | 0.01 | NA | NA | NA | NA | NA | NA | NA | NA | NA | NA | NA | NA |
| CR1 | NA | NA | NA | NA | rs6656401 | 1.38 | 9.02E-03 | 0.04 | NA | NA | NA | NA | rs6733839 | 1.2 | 2.10E-44 | 0.407 |
| CYB561 | NA | NA | NA | NA | NA | NA | NA | NA | NA | NA | NA | NA | rs138190086 | 1.3 | 5.30E-09 | 0.02 |
| CYP27C1 | rs7585314 | 0.75 | 3.00E-03 | 0.33 | NA | NA | NA | NA | NA | NA | NA | NA | NA | NA | NA | NA |
| DSG2 | NA | NA | NA | NA | NA | NA | NA | NA | NA | NA | NA | NA | rs8093731 | 0.54 | 4.60E-08 | 0.017 |
| ECHDC3 | NA | NA | NA | NA | NA | NA | NA | NA | NA | NA | NA | NA | rs7920721 | 1.08 | 1.80E-11 | 0.39 |
| EED | NA | NA | NA | NA | rs3851179 | 0.8 | 1.71E-05 | 0.39 | NA | NA | NA | NA | rs4844610 | 1.17 | 3.60E-24 | 0.187 |
| EFCAB14 | NA | NA | NA | NA | rs7519866 | 0.71 | 9.70E-06 | 0.37 | NA | NA | NA | NA | NA | NA | NA | NA |
| ENOX1 | rs17460623 | 0.49 | 9.37E-06 | 0.1 | NA | NA | NA | NA | NA | NA | NA | NA | NA | NA | NA | NA |
| EPHA1 | NA | NA | NA | NA | NA | NA | NA | NA | NA | NA | NA | NA | rs10808026 | 0.9 | 1.30E-10 | 0.199 |
| FBXL7 | NA | NA | NA | NA | NA | NA | NA | NA | rs75002042 | 0.6 | 4.70E-09 | 0.08 | NA | NA | NA | NA |
| FERMT2 | NA | NA | NA | NA | NA | NA | NA | NA | NA | NA | NA | NA | rs17125924 | 1.14 | 1.40E-09 | 0.093 |
| FST | NA | NA | NA | NA | NA | NA | NA | NA | NA | NA | NA | NA | rs35868327 | 0.68 | 2.60E-07 | 0.013 |
| GATA3 | NA | NA | NA | NA | rs1273007 | 0.68 | 3.08E-06 | 0.27 | NA | NA | NA | NA | NA | NA | NA | NA |
| GPR141 | NA | NA | NA | NA | NA | NA | NA | NA | NA | NA | NA | NA | rs4723711 | 0.94 | 2.80E-07 | 0.356 |
| GRIA4 | NA | NA | NA | NA | NA | NA | NA | NA | NA | NA | NA | NA | rs509512 | 0.75 | 7.37E-06 | 0.43 |
| GRIN3B | rs115882880 | 1.55 | 6.34E-08 | 0.11 | NA | NA | NA | NA | NA | NA | NA | NA | NA | NA | NA | NA |
| HAS2 | rs956225 | 0.3 | 8.71E-06 | 0.03 | NA | NA | NA | NA | NA | NA | NA | NA | NA | NA | NA | NA |
| HBEGF | NA | NA | NA | NA | NA | NA | NA | NA | NA | NA | NA | NA | rs11168036 | 1.12 | 3.20E-07 | 0.5 |
| HID1 | NA | NA | NA | NA | NA | NA | NA | NA | NA | NA | NA | NA | rs71380849 | 1.47 | 9.10E-07 | 0.06 |
| HLA-DQA1 | NA | NA | NA | NA | NA | NA | NA | NA | NA | NA | NA | NA | rs9271192 | 1.11 | 1.60E-08 | 0.276 |
| HLA-DRB1 | NA | NA | NA | NA | NA | NA | NA | NA | NA | NA | NA | NA | rs9271058 | 1.1 | 1.40E-11 | 0.27 |
| IGLON5 | rs10419982 | 1.38 | 5.40E-04 | 0.4 | NA | NA | NA | NA | NA | NA | NA | NA | NA | NA | NA | NA |
| INPP5D | NA | NA | NA | NA | NA | NA | NA | NA | NA | NA | NA | NA | rs10933431 | 0.91 | 3.40E-09 | 0.223 |
| IQCK | NA | NA | NA | NA | NA | NA | NA | NA | NA | NA | NA | NA | rs7185636 | 0.92 | 2.40E-08 | 0.18 |
| KAZN | NA | NA | NA | NA | NA | NA | NA | NA | NA | NA | NA | NA | rs7527934 | 0.87 | 5.87E-06 | 0.257 |
| LTBP2 | NA | NA | NA | NA | NA | NA | NA | NA | NA | NA | NA | NA | rs2043948 | 1.27 | 4.44E-07 | 0.077 |
| MARK4 | NA | NA | NA | NA | rs597668 | 0.88 | 8.23E-03 | 0.43 | NA | NA | NA | NA | rs597668 | 1.17 | 6.45E-09 | 0.154 |
| MEF2C | NA | NA | NA | NA | NA | NA | NA | NA | NA | NA | NA | NA | rs190982 | 0.92 | 2.50E-06 | 0.408 |
| MS4A4A | NA | NA | NA | NA | NA | NA | NA | NA | NA | NA | NA | NA | rs4938933 | 0.88 | 1.66E-09 | 0.396 |
| MS4A6A | NA | NA | NA | NA | NA | NA | NA | NA | NA | NA | NA | NA | rs7933202 | 0.89 | 1.90E-19 | 0.391 |
| MSX2 | rs145848414 | 2.29 | 6.90E-08 | 0.04 | NA | NA | NA | NA | NA | NA | NA | NA | NA | NA | NA | NA |
| MTHFD1L | NA | NA | NA | NA | NA | NA | NA | NA | NA | NA | NA | NA | rs11754661 | 2.1 | 1.90E-10 | 0.07 |
| NDUFAF6 | NA | NA | NA | NA | NA | NA | NA | NA | NA | NA | NA | NA | rs4735340 | 0.94 | 9.20E-08 | 0.476 |
| NECTIN2 | NA | NA | NA | NA | NA | NA | NA | NA | NA | NA | NA | NA | rs6859 | 1.41 | 9.60E-08 | 0.46 |
| NYAP1 | NA | NA | NA | NA | NA | NA | NA | NA | NA | NA | NA | NA | rs12539172 | 0.92 | 9.30E-10 | 0.303 |
| OARD1 | NA | NA | NA | NA | NA | NA | NA | NA | NA | NA | NA | NA | rs114812713 | 1.32 | 2.10E-13 | 0.03 |
| PALM2AKAP2 | NA | NA | NA | NA | rs913360 | 1.56 | 1.83E-07 | 0.28 | NA | NA | NA | NA | NA | NA | NA | NA |
| PGM2L1 | rs3888908 | 1.72 | 9.52E-06 | 0.15 | NA | NA | NA | NA | NA | NA | NA | NA | NA | NA | NA | NA |
| PICALM | rs12795381 | 0.49 | 8.60E-03 | 0.04 | NA | NA | NA | NA | NA | NA | NA | NA | rs10792832 | 0.88 | 6.50E-16 | 0.358 |
| POLN | rs1923775 | 1.6 | 5.61E-06 | 0.25 | NA | NA | NA | NA | NA | NA | NA | NA | NA | NA | NA | NA |
| PROX1 | rs340849 | 0.59 | 7.52E-06 | 0.2 | NA | NA | NA | NA | NA | NA | NA | NA | NA | NA | NA | NA |
| PTK2B | NA | NA | NA | NA | NA | NA | NA | NA | NA | NA | NA | NA | rs73223431 | 1.1 | 6.30E-14 | 0.367 |
| RAD51AP2 | rs11889338 | 1.55 | 8.94E-06 | 0.26 | NA | NA | NA | NA | NA | NA | NA | NA | NA | NA | NA | NA |
| RDH13 | NA | NA | NA | NA | NA | NA | NA | NA | NA | NA | NA | NA | rs6509916 | 1.34 | 5.83E-06 | 0.46 |
| RIN3 | NA | NA | NA | NA | rs11621843 | 1.47 | 5.19E-06 | 0.26 | NA | NA | NA | NA | NA | NA | NA | NA |
| RNF6 | rs17511627 | 1.75 | 5.01E-06 | 0.17 | NA | NA | NA | NA | NA | NA | NA | NA | NA | NA | NA | NA |
| SLC10A2 | rs16961023 | 2.77 | 8.01E-07 | 0.02 | NA | NA | NA | NA | NA | NA | NA | NA | NA | NA | NA | NA |
| SLC24A4 | NA | NA | NA | NA | NA | NA | NA | NA | NA | NA | NA | NA | rs12881735 | 0.92 | 7.40E-09 | 0.221 |
| SLC4A1AP | rs17006206 | 2.05 | 2.30E-06 | 0.1 | NA | NA | NA | NA | NA | NA | NA | NA | NA | NA | NA | NA |
| SORL1 | NA | NA | NA | NA | rs3781834 | 0.74 | 7.30E-07 | 0.23 | NA | NA | NA | NA | rs3851179 | 0.88 | 6.00E-25 | 0.356 |
| SPI1 | NA | NA | NA | NA | NA | NA | NA | NA | NA | NA | NA | NA | rs3740688 | 0.92 | 5.40E-13 | 0.448 |
| SPPL2A | NA | NA | NA | NA | NA | NA | NA | NA | NA | NA | NA | NA | rs10467994 | 0.94 | 3.90E-07 | 0.333 |
| STK24 | rs912330 | 0.54 | 3.79E-06 | 0.14 | NA | NA | NA | NA | NA | NA | NA | NA | NA | NA | NA | NA |
| TAS2R60 | rs4595035 | 1.25 | 9.40E-03 | 0.43 | NA | NA | NA | NA | NA | NA | NA | NA | NA | NA | NA | NA |
| TBX3 | rs10850408 | 0.63 | 9.25E-07 | 0.34 | NA | NA | NA | NA | NA | NA | NA | NA | NA | NA | NA | NA |
| TMPRSS15 | NA | NA | NA | NA | NA | NA | NA | NA | NA | NA | NA | NA | rs2825544 | 1.14 | 4.85E-07 | 0.346 |
| TMTC1 | rs302318 | 0.59 | 1.97E-06 | 0.26 | NA | NA | NA | NA | NA | NA | NA | NA | NA | NA | NA | NA |
| TNFAIP3 | NA | NA | NA | NA | NA | NA | NA | NA | NA | NA | NA | NA | rs679670 | 0.74 | 9.83E-06 | 0.37 |
| TREM2 | NA | NA | NA | NA | NA | NA | NA | NA | NA | NA | NA | NA | rs75932628 | 2.08 | 2.70E-15 | 0.008 |
| TRIP4 | NA | NA | NA | NA | NA | NA | NA | NA | NA | NA | NA | NA | rs74615166 | 1.519 | 3.27E-03 | 0.023 |
| TSPOAP1 | NA | NA | NA | NA | NA | NA | NA | NA | NA | NA | NA | NA | rs2632516 | 0.94 | 5.30E-08 | 0.44 |
| WWOX | NA | NA | NA | NA | NA | NA | NA | NA | NA | NA | NA | NA | rs62039712 | 1.16 | 3.70E-08 | 0.116 |
| ZC3H3 | rs3750208 | 0.37 | 7.28E-06 | 0.04 | NA | NA | NA | NA | NA | NA | NA | NA | NA | NA | NA | NA |
| ZCWPW1 | NA | NA | NA | NA | NA | NA | NA | NA | NA | NA | NA | NA | rs1476679 | 0.92 | 7.40E-06 | 0.287 |

**Supplementary Figure 1** – Classification of studies by the analysis type and stage of analyses

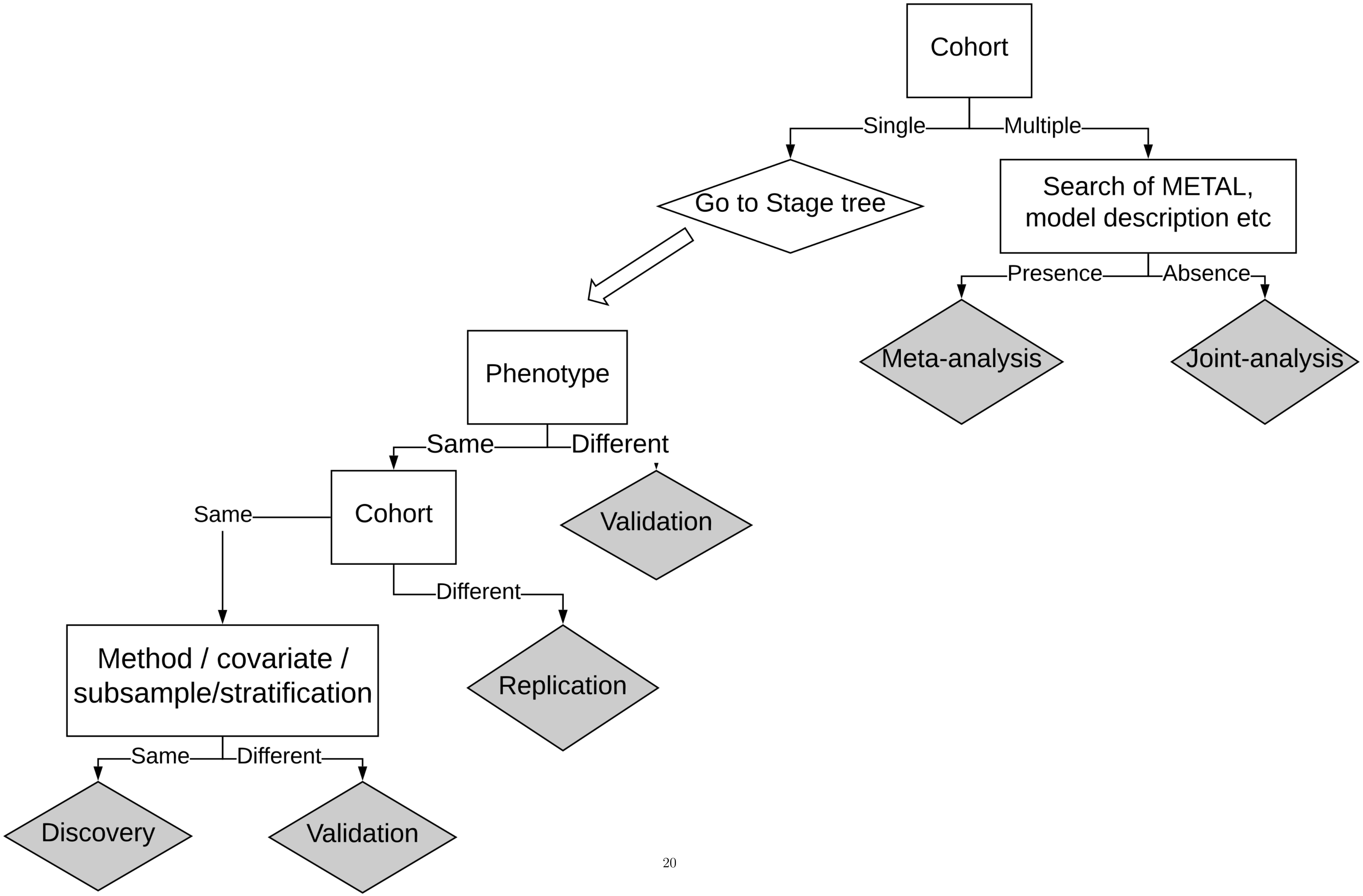
